# Post-viral parenchymal lung disease following COVID-19 and viral pneumonitis hospitalisation: A systematic review and meta-analysis

**DOI:** 10.1101/2021.03.15.21253593

**Authors:** Laura Fabbri, Samuel Moss, Fasihul Khan, Wenjie Chi, Jun Xia, Karen A. Robinson, Alan Smyth, Gisli Jenkins, Iain Stewart

**Affiliations:** Division of Respiratory Medicine, University of Nottingham, Nottingham, UK; Nottingham Biomedical Research Centre, National Institute for Health Research, UK; National Heart and Lung Institute, Imperial College London, London, UK; Systematic Review Solutions Ltd, Ingenuity Centre, Nottingham, UK; Institute of Mental Health, University of Nottingham, Nottingham, UK; Department of Medicine, Johns Hopkins School of Medicine, Johns Hopkins University, US; Division of Child Health, Obstetrics & Gynaecology, University of Nottingham, Nottingham, UK

**Author notes:** Correspondence to: Dr Iain Stewart, Guy Scadding Building, Imperial College London, SW3 6LY. **Funding:** National Institute for Health Research (NIHR). **Summary competing interests:** LF, SM, FK, WC, JX, KR, AS and IS report no competing interests relating to the manuscript, GJ reports NIHR BRC salaries, studentships, professorship (RP-2017-08-ST2-014). **Declarations:** this manuscript is available on a preprint server, DOI: 10.1101/2021.03.15.21253593.

**Keywords:** COVID19, CT, gas transfer, lung sequelae, restrictive impairment, pulmonary fibrosis

## Abstract

**Background:** Approximately half of COVID-19 survivors present persisting breathlessness, which may include development of pulmonary fibrosis.

**Research Question:** What is the prevalence of long-term radiological and functional pulmonary sequelae of parenchymal lung disease following hospitalisation with COVID-19 and other viral pneumonia?

**Study design and methods:** We performed systematic review and random effects meta-analysis of studies in adults hospitalised with SARS-CoV-2, SARS-CoV, MERS-CoV, or Influenza pneumonia and followed within 12 months from discharge. Searches were run on MEDLINE and Embase, updated 29 July 2021. Primary outcomes were proportion of 1) radiologic sequelae at CT scans; 2) restrictive impairment; 3) impaired gas transfer. Heterogeneity was explored in meta-regression.

**Results:** Ninety-five studies were included for qualitative synthesis, of which 70 were suitable for meta-analysis, including 60 studies of SARS-CoV-2 with a median follow up of 3 months. In SARS-CoV-2 the overall estimated proportion of inflammatory changes during follow up was 0.50 (95%CI 0.41 to 0.58, I^2^=94.6%), whilst fibrotic changes were estimated at 0.29 (95%CI 0.22 to 0.37, I^2^=94.1%). Inflammatory changes reduced compared with CTs performed during hospitalisation (−0.47; 95%CI -0.56 to -0.37), whereas no significant resolution was observed in fibrotic changes (−0.09; 95%CI -0.25 to 0.07). Impaired gas transfer was estimated at 0.38 (95%CI 0.32 to 0.44, I^2^=92.1%), which was greater than estimated restrictive impairment (0.17; 95%CI 0.13 to 0.23, I^2^=92.5%). High heterogeneity means that estimates should be interpreted with caution. Confidence in the estimates was deemed low due to the heterogeneity and because studies were largely observational without controls.

**Interpretation:** A substantial proportion of radiological and functional sequelae consistent with parenchymal lung disease are observed following COVID-19 and other viral pneumonitis. Estimates of prevalence are limited by differences in case mix and initial severity. This highlights the importance of extended radiological and functional follow-up post hospitalisation.

**PROSPERO registration:** CRD42020183139 (April 2020)

## Introduction

Since COVID-19, the disease caused by SARS-CoV-2, was declared a global pandemic,^1^ over 220 million individuals have been infected (September 2021).^2^ The clinical spectrum of COVID-19 is wide, and can range from asymptomatic or mild flu-like symptoms, to severe viral pneumonia, requiring hospital admission, oxygen administration, and mechanical ventilation.^3^ Emerging data suggest that approximately half of COVID-19 survivors experience a long-term multisystemic syndrome characterized by chronic breathlessness and other abnormalities.^4-6^ The causes for the persistent respiratory symptoms have not been clearly elucidated, however, post-mortem studies on COVID-19 patients have highlighted diffuse parenchymal alterations, with alveolar damage, exudation, and development of pulmonary fibrosis.^7-9^

Pulmonary fibrosis is characterised by a dysregulated remodelling of the lung parenchyma. It can occur after a lung injury, although the cause cannot always be identified. Viral agents are considered important insults, with scientific rationale to implicate their role in fibrosis pathogenesis, although empirical evidence that suggests they can promote chronic parenchymal lung disease is limited.^10 11^ Fibrotic sequelae have been highlighted in the follow up of SARS-CoV and MERS-CoV.^12-14^ Similarly, Influenza viruses have also been proposed to promote the development of pulmonary fibrosis.^15,16^

Given the exceptional rate of COVID-19 spread and the longer-term impact on quality of life, particularly breathlessness, it is possible that parenchymal lung abnormalities may be a long-term consequence in survivors. We undertook a systematic review and meta-analysis to assess the prevalence of lung sequelae in people hospitalised with viral pneumonitis, focusing on CT scans and pulmonary function tests as non-invasive diagnostic exams routinely used to assess the presence of lung abnormalities. ^17,18^

## Methods

### Search strategy and selection criteria

The systematic review and meta-analysis was conducted in accordance with a protocol registered with the International Prospective Register of Systematic Reviews (PROSPERO) on 30^th^ April 2020 (registration number CRD42020183139). The review has been reported following PRISMA and PICO guidelines.^19,20^ All original research reporting outcomes in populations of hospitalized adult patients (aged >18) with presumed or confirmed viral infection by SARS-CoV-2, SARS-CoV, MERS-CoV or Influenza viruses were considered eligible for inclusion. No intervention was assessed relative to a control group. Comparisons were made between radiological sequelae types and metrics of lung function impairment, and compared with findings during hospitalisation where available. The pre-specified primary outcomes within 12 months of hospitalisation were: 1) presence of radiologic sequelae at follow-up CT scans; 2) presence of restrictive lung function impairment; 3) presence of reduced diffusing capacity for carbon monoxide (DL_CO_). Inflammatory radiological findings were defined as ground-glass opacification or consolidation. Fibrotic radiological findings were defined as either reticulation, lung architectural distortion, interlobular septal thickening, traction bronchiectasis, or honeycombing. Restrictive lung impairment was defined as a total lung capacity (TLC) <80% predicted value or forced vital capacity (FVC) < 80% predicted value with normal-to-high FEV1/FVC ratio. Impaired gas transfer was defined as percent predicted DL_CO_ < 80%.

Searches were performed in MEDLINE (1946 to latest), Embase (1974 to latest), and Google Scholar. Hand searches were conducted of the reference lists of eligible primary studies, and relevant review articles. No language criteria were applied. Pre-prints, abstracts, and non-original studies were excluded. Searches were last updated on 29^th^ of July 2021. Searches were carried out using patient-related, treatment-related, and outcomes-related terms (Supplementary Figure 1). Titles and abstracts were screened in duplicate, followed by full-text review. Disagreements between reviewers were resolved by consensus with a third reviewer.

### Data analysis

Data from the selected articles were extracted independently using a proforma by reviewers and mutually confirmed. Extracted data included study design, viral agent, methods of diagnosis, participant demographics, severity of acute infection (ventilatory requirements), as well as CT and lung function outcomes. Baseline investigations were defined as those performed during hospitalization, and follow-up as obtained after discharge; baseline data were only extracted where studies reported follow-up. If more than one follow-up visit was reported, the most complete sample size followed by the latest examination within 12 months from discharge was extracted in a hierarchical manner. Where data were not reported in the text, we contacted corresponding authors. Absolute values of the number of people meeting outcome criteria and number of people with exam results available were extracted as numerator and denominator, respectively.

Meta-analyses of proportions were performed where sufficient studies reported data, enabling an estimation of the prevalence of outcomes. Cohorts with fewer than ten cases (SARS-CoV, Influenza) or 25 cases (SARS-CoV-2) were excluded from quantitative synthesis owing to risk of selection bias when estimating proportions. Separate analyses were performed in each viral subtype (SARS-CoV-2, SARS-CoV, Influenza) and according to the type of radiological (inflammatory, fibrotic) or physiological (restrictive impairment, impaired gas transfer) outcome. Quantitative synthesis and random effects meta-analysis were performed in Stata SE16 (TX: StataCorp LLC) using the *metaprop* command, which computes 95% confidence intervals based on binomial distribution and applies the Freeman-Tukey double arcsine transformation to support inclusion of observations of 0% and 100%.^21^ Heterogeneity was assessed with I^2^; we report all estimates regardless of heterogeneity.

For SARS-CoV-2 studies, meta-regression was performed to assess the residual heterogeneity after adjustment for key study characteristics, timing of follow-up (months), severity of cohort (mild, moderate, severe), prospective design, evidence of selection bias (strict inclusion criteria based on indication for CT or where less than 60% of screened patients tested for outcomes), and approach to radiological classification (study author defined, or by review). Residual heterogeneity is assessed with I^2^, R^2^ is used to describe the variance in estimate explained by adjusted models. Reliability of estimates was assessed through sensitivity analysis in a restricted timeframe of 3-6 months follow up, and in sub analysis on studies that reported baseline quantifications.

The risk of bias in individual studies was assessed by two authors independently using appropriate assessment tools available from the CLARITY Group at McMaster University,^22^ through criteria specific for study design. We assessed exposure, the outcomes of interest, prognostic factors, interventions, adequacy of follow-up, and co-interventions. Randomized controlled trials were evaluated on random sequence generation, allocation concealment, blinding, adequacy of follow up, selective reporting, and other possible causes of risks of bias. Any disagreements were resolved by consensus with a third reviewer.

The quality of the evidence for each overall estimate of proportion was evaluated using the GRADE guidance.^24^ Retrospective observational studies were considered very low but could be upgraded, whilst prospective randomised studies were deemed high and could be downgraded. Analytical and publication risks of bias, inconsistency, indirectness, and imprecision in reporting were assessed. An overall judgement of ‘high’, ‘moderate’, ‘low’, or ‘very low’ was provided for the quality of the cumulative evidence for review outcomes.

## Results

A total of 8321 records were identified from databases and hand searches. After title and abstract screening, 131 unique full-text manuscripts were assessed for eligibility, and 95 were included for qualitative synthesis (89 in English, 6 in Chinese). A total of 70 studies were included in the quantitative synthesis (Figure 1). Among the manuscripts included, 60 reported infections by SARS-CoV-2;^4,23,25-83^ 18 by SARS-CoV;^13,14,84-100^ 1 by MERS-CoV;^101^ 16 by Influenza (11 subtype H1N1, 1 subtype H5N1, 1 subtype H3N2, 2 subtype H7N9 and 1 study both H1N1 and H7N9).^102-117^ All studies were observational in design, with the exception of a single randomized control trial.^97^ We focus reporting on changes subsequent to a SARS-CoV-2 infection, quantitative synthesis for SARS-CoV and influenza are provided in supplemental material.

**Figure 1:**
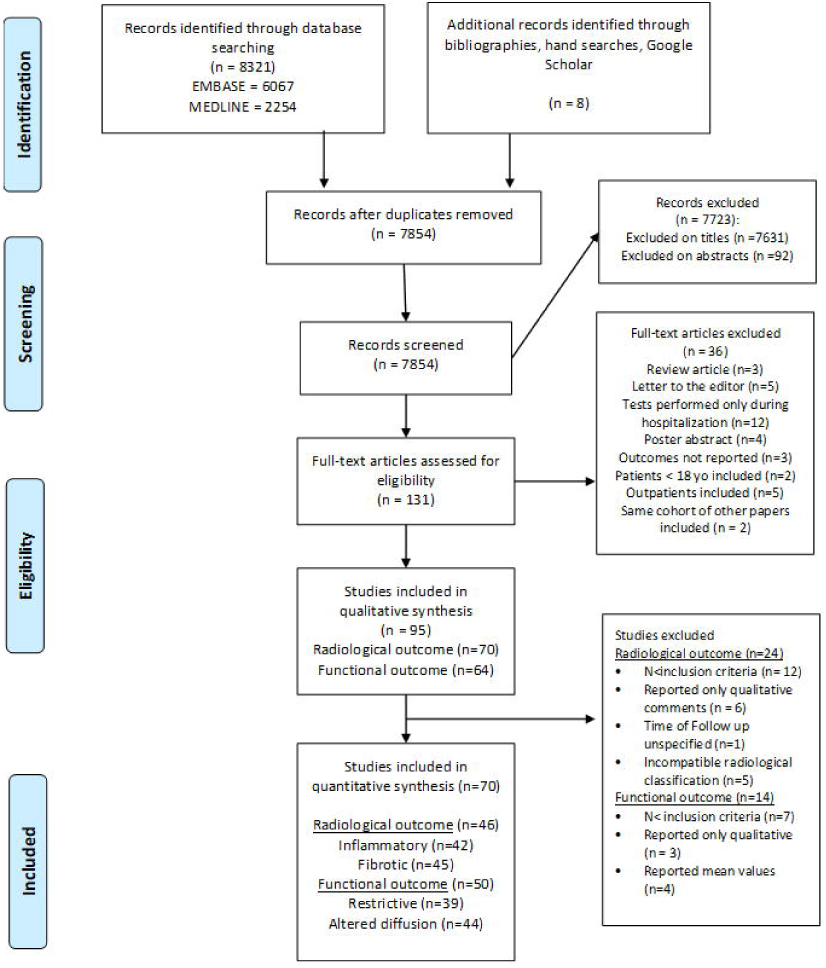
Systematic search and screening strategy. Flow diagram illustrates systematic search and screening strategy, including numbers meeting eligibility criteria and numbers excluded. Searches updated on the 29^th^ of July 2021.

Individual SARS-CoV-2 study characteristics are presented in Table 1, and supplementary Table 1. Risk of bias assessment identified a number of limitations and possible causes of biases. A total of ten studies did not specify whether any serological or molecular testing was performed or referred to national guidelines at the time the study was conducted. Inclusion and exclusion criteria differed among studies, indicating that the severity of patients enrolled and care pathways followed may represent a possible selection bias. Few studies investigated the presence of previous respiratory diseases or considered it as an exclusion criteria, others were restricted to include only symptomatic patients or perform follow-up CT where there was a clinical indication, such as abnormalities on chest X-Ray (CXR) or reduced DL_CO_.^14,41,59,89^ Details for all the studies are presented in Supplementary Tables 1-2; Supplementary Figure 2a -2b.

**Table 1.**
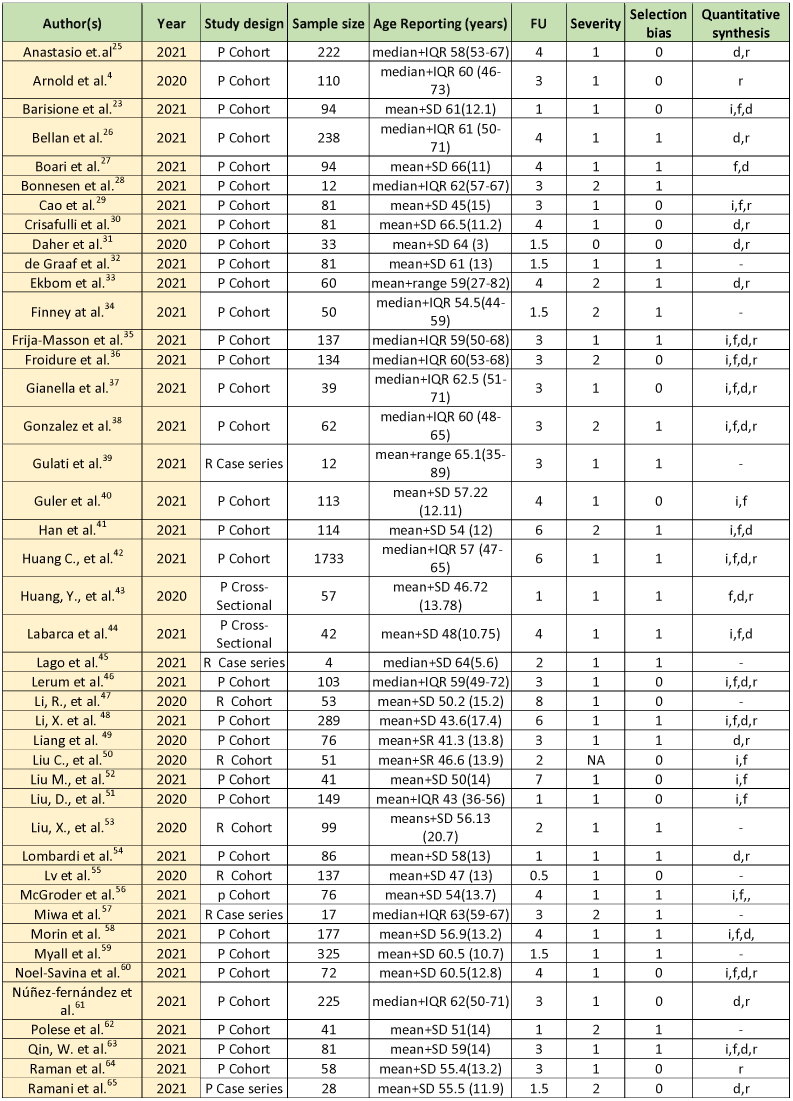
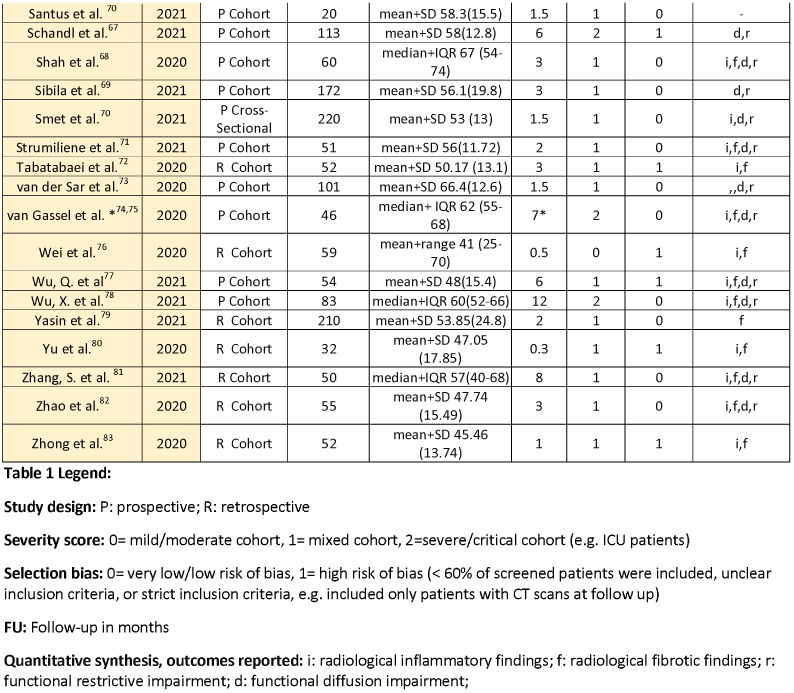
SARS-CoV-2 Studies overview.

A total of 70 studies described thoracic CT findings, 46 were included in meta-analysis of radiological outcome of SARS-CoV-2. Causes of exclusion are listed in Figure 1. The median follow-up time was 3 months. Within 12 months following hospitalisation for SARS-CoV-2 infection, the overall estimated proportion of chest CT inflammatory changes was 0.50 (95% CI 0.41 to 0.58. I^2^= 95.0%) on a total of 2670 CT scans, whilst fibrotic changes had an estimated proportion of 0.29 (95% CI 0.22 to 0.37. I^2^=94.1%) assessed on 2811 exams. Severe heterogeneity was observed in overall estimates (Figure 2). In meta-regression, adjustment for timing of follow-up significantly reduced the residual heterogeneity in overall estimate of inflammatory changes to 73.1% (Supplementary Table 4). Adjustment for timing reduced residual heterogeneity in the overall estimate of fibrotic change to 70.3%. No other characteristics were observed to significantly contribute to heterogeneity, including severity of cohort, prospective design or risk of selection bias (Supplementary Tables 4-5).

**Figure 2.**
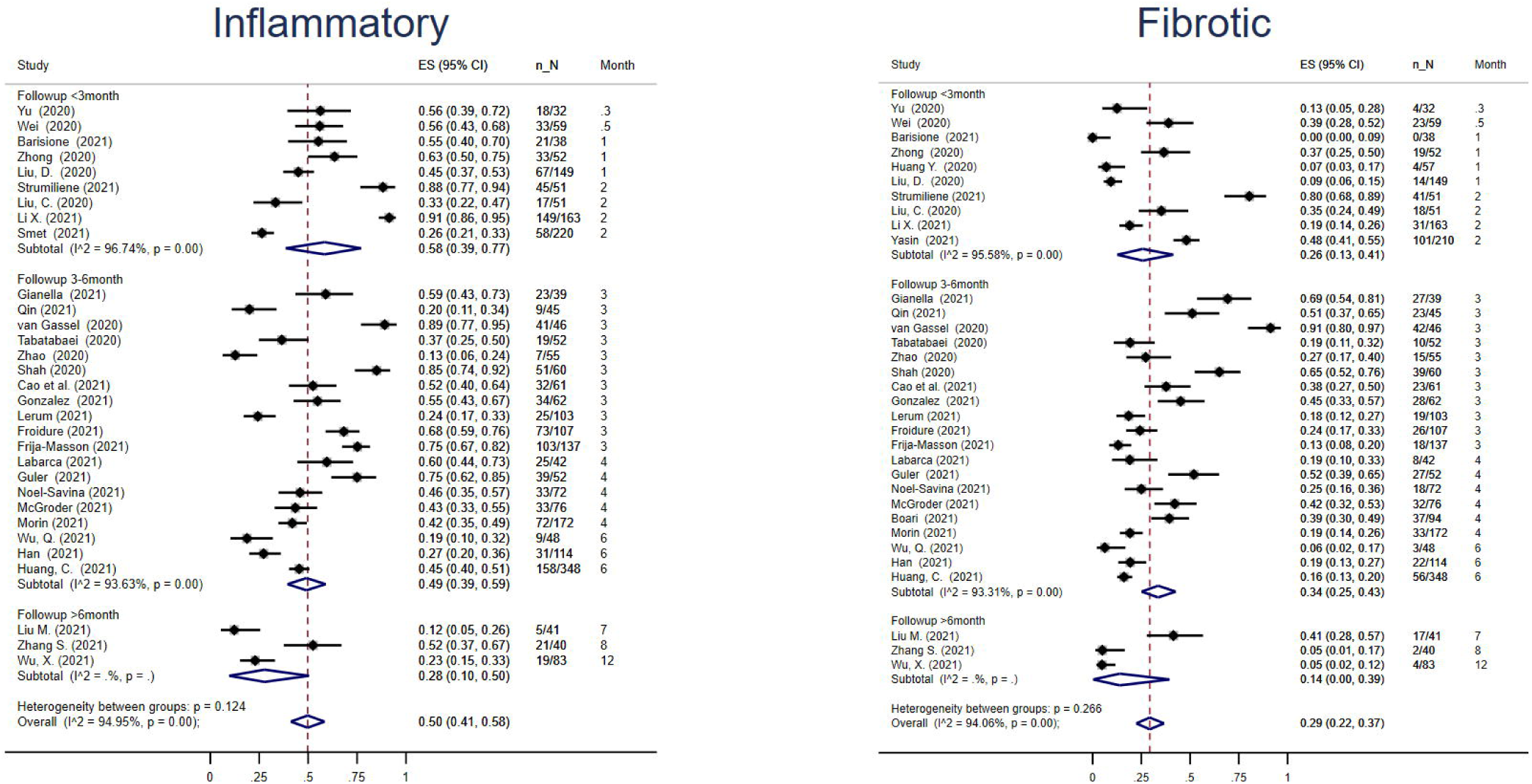
Radiological findings at follow-up in SARS-CoV-2 studies. Estimates are reported as proportion of number of CT scans showing the outcome of interest (n) on the total number of exams performed (N) and 95% confidence interval. Inflammatory radiological findings were defined as ground-glass opacification or consolidation. Fibrotic radiological findings were defined as either reticulation, lung architectural distortion, interlobular septal thickening, traction bronchiectasis, or honeycombing.

We subsequently selected a restricted follow up timeframe of 3-6 months to minimise inconsistency in follow-up time (Supplementary Figure 3). Within this sensitivity analysis we observed similar estimates. The overall estimated proportion of chest CT inflammatory changes was 0.49 (95%CI 0.39 to 0.59, I^2^=93.6%), whilst residual heterogeneity reduced to 71.0% after adjustment for timing of follow-up, explaining 9.3% of the variance. Prospective design and the severity of the cohort also contributed to variance in the estimate: R^2^ 11.7% and 2.6%, respectively (Supplementary Table 4). The overall estimated proportion of fibrotic changes was 0.34 (95%CI 0.25 to 0.43, I^2^=93.3%), adjustment for timing in meta regression reduced residual heterogeneity to 63.7%, explaining 21.0% of variance, whilst risk of selection bias and approach to radiological classification also contributed to variance in the estimate: R^2^ 21.1% and 2.9%, respectively (Supplementary Table 5). The lowest unadjusted heterogeneity in estimate was observed at the 4-month follow-up, where inflammatory changes were estimated at a proportion of 0.53 (95%CI 0.41 to 0.64, I^2^=81.4%) whilst fibrotic changes were estimated at 0.32 (95%CI 0.22 to 0.43, I^2^=84.9%) (Supplementary Figure 4).

In sub analysis of studies that reported baseline outcomes, estimates of inflammatory changes were 0.92 (95%CI 0.87 to 0.96, I^2^=89.4%) at baseline, 0.44 (95%CI 0.35 to 0.53, I^2^=89.3%) at follow-up, resulting in an estimated difference in proportion of -0.47 (−0.56 to -0.37, I^2^=87.8%) over time (Figure 3). Estimates of fibrotic changes at baseline were 0.32 (95%CI 0.15 to 0.52, I^2^=98.0%) and 0.26 (95%CI 0.17 to 0.36, I^2^=92.9%) at follow-up, with an estimated difference in proportion of -0.09 (95%CI -0.25 to 0.07, I^2^=96.4%) over time (Figure 4). After adjustment for timing of follow-up, overall heterogeneity of estimates of inflammatory changes at matched follow up were reduced from 89.3% to 55.7%, whilst heterogeneity in estimates of fibrotic changes at matched follow up were reduced from 92.9% to 58.3% (Supplementary Tables 4-5; Supplementary Figure 4). Prospective design contributed 5.3% of variance in estimates of inflammatory changes, whilst selection bias explained 8.3% of variance in estimates of fibrotic changes.

**Figure 3.**
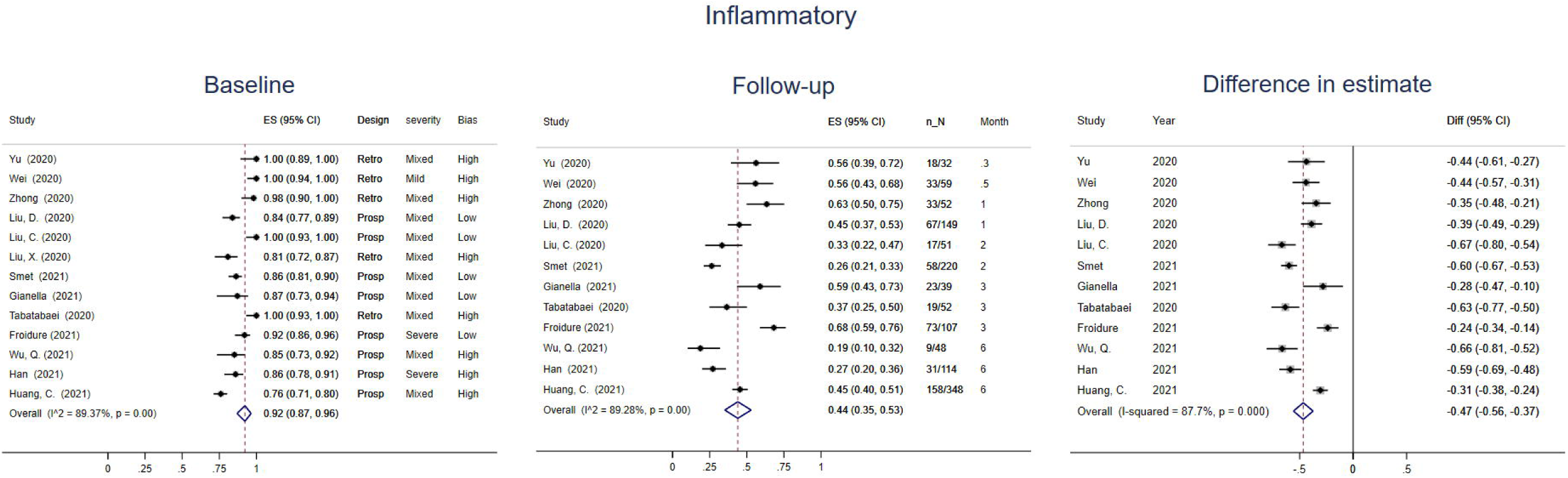
Inflammatory findings at baseline and matched follow-up in SARS-CoV-2 studies: sub analysis. Estimates are reported as proportion of number of CT scans showing the outcome of interest (n) on the total number of exams performed (N) and 95% confidence interval. Baseline defined as during hospitalisation, follow-up defined as post discharge. Inflammatory radiological findings were defined as ground-glass opacification or consolidation.

**Figure 4.**
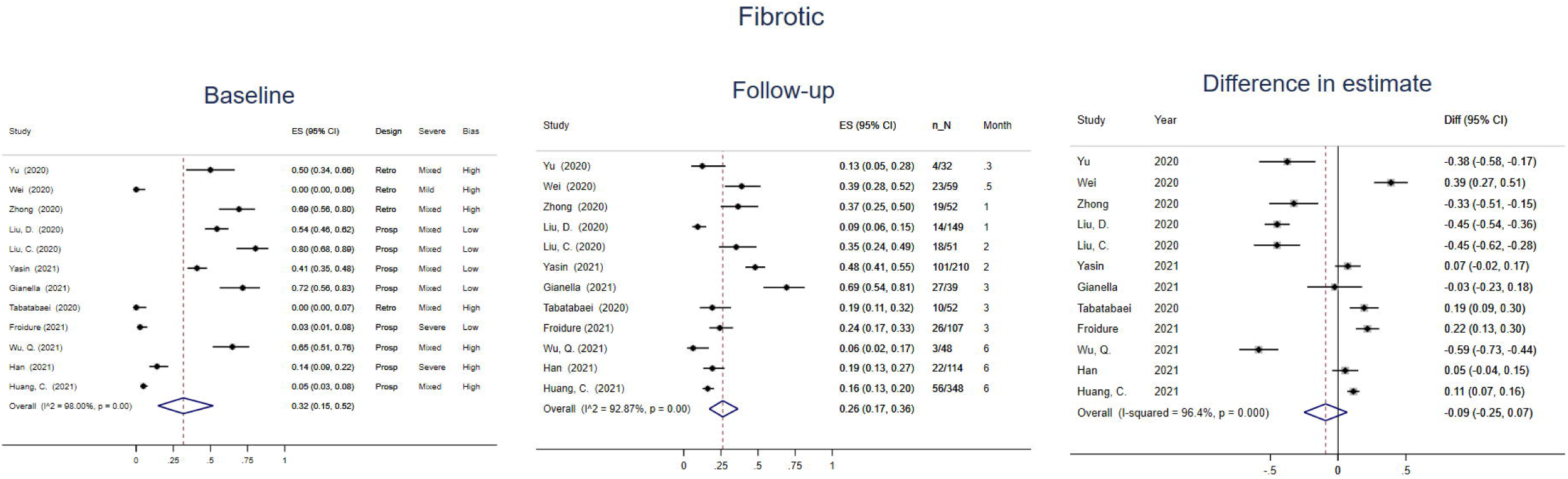
Radiological fibrotic findings at baseline and matched follow-up in SARS-CoV-2 studies: sub analysis. Estimates are reported as proportion of number of CT scans showing the outcome of interest (n) on the total number of exams performed (N) and 95% confidence interval. Baseline defined as during hospitalisation, follow-up defined as post discharge. Fibrotic radiological findings were defined as either reticulation, lung architectural distortion, interlobular septal thickening, traction bronchiectasis, or honeycombing.

In separate viral agent strata, the estimated proportion of patients with inflammatory changes during follow-up CT scans was 0.81 (95%CI 0.58 to 0.97, I^2^=91.8%), and 0.61 (95%CI 0.27 to 0.90, I^2^=93.3%) following SARS-CoV and Influenza infections, respectively. The overall estimate of fibrotic changes during follow-up was 0.66 (95%CI 0.43 to 0.86, I^2^=92.8%), and 0.27 (95%CI 0.15 to 0.40. I^2^= 57.1%) following SARS-CoV and Influenza infections, respectively (Supplementary Figure 5).

Lung function sequelae were described in a total of 64 papers, with 50 reaching sample size criteria for inclusion in quantitative synthesis. A total of 3146 tests for restrictive impairment and 3419 for impaired DLco were included following SARS-CoV-2 infection. Follow-up lung function tests were performed at a median of 3 months after discharge. The overall estimated proportion of individuals with impaired gas transfer during follow-up was 0.38 (95% CI 0.32 to 0·44. I^2^=92.1%) and 0.38 (95%CI 0.32 to 0.44, I^2^=91.2%) in the restricted 3-6 months time frame (Figure 5; Supplementary Figure 6). In meta regression, adjustment for timing of follow-up reduced residual heterogeneity to 63.2% and 59.4% for impaired gas transfer overall and when restricted to 3-6 months window, respectively (Supplementary Table 6; Supplementary Figure 9). The overall estimated proportion of individuals with restrictive impairment within 12 months was 0.17 (95% CI 0.13 to 0.23. I^2^=92.5%) and 0.14 (95%CI 0.10 to 0.19, I^2^=86.6%) when follow up was restricted to 3-6 months (Figure 5; Supplementary Figure 7). In meta regression, adjustment for timing of follow-up reduced residual heterogeneity to 60.1% and 16.7% for restrictive impairment overall and when restricted to 3-6 months follow-up, respectively (Supplementary Table 7; Supplementary Figure 9). Severity of the cohort explained 35.5% of the variance in estimate in sensitivity analysis.

**Figure 5.**
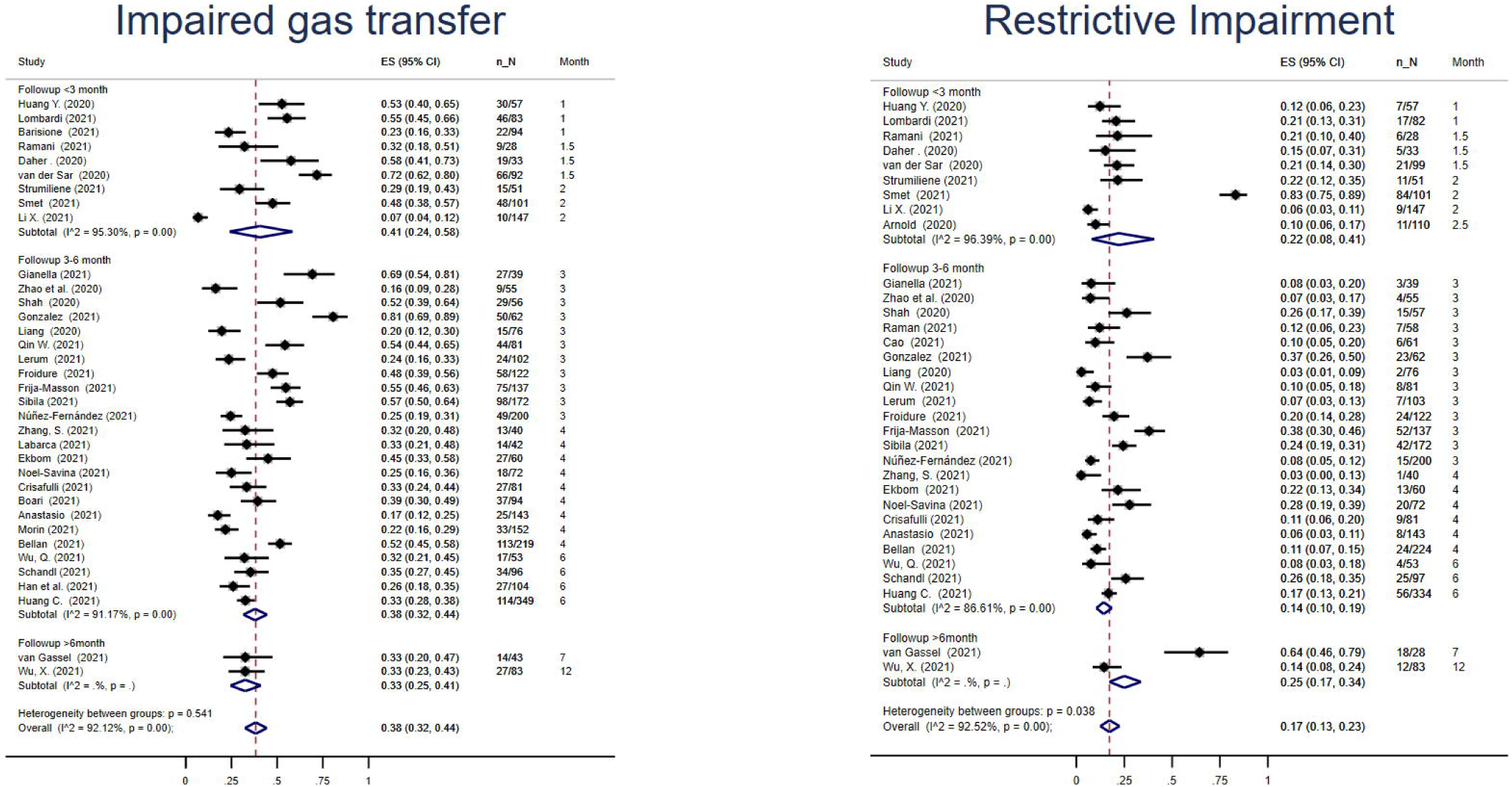
Pulmonary function testing at follow-up. Estimates are reported as proportion of number of tests showing the outcome of interest (n) on the total number of exams performed (N) and 95% confidence interval. Restrictive lung impairment was defined as a total lung capacity (TLC) <80% predicted value or forced vital capacity (FVC) < 80% predicted value with normal-to-high FEV1/FVC ratio. Impaired gas transfer was defined as percent predicted DLCO < 80%.

Estimates of the prevalence of restrictive impairment were similarly low across other viral pneumonias, 0.10 (95%CI 0.05 to 0.17, I^2^=80.2%) for SARS-CoV and in 6/73 participants with MERS-CoV (Supplementary Figure 7). Estimates of the prevalence of impaired gas transfer were similar in SARS-CoV compared to SARS-CoV-2 (0.36; 95%CI 0.27 to 0.46, I^2^=84.4%), whilst prevalence was estimated to be higher following influenza (0.54; 95%1I 0.43 to 0.65), and a single study of MERS-CoV identified gas transfer impairments in 25/73 participants.

Based on the GRADE framework, we have low confidence in estimates for all outcomes. All studies included in the quantitative synthesis had an observational design and moderate risk of bias as possible confounding factors were not extensively assessed and could not be modelled in estimates of proportion. Inconsistency between studies was considered moderate due to the substantial heterogeneity that could be reduced by adjustment for timing. No causes of indirectness were detected since all study subjects had confirmed viral pneumonia, although severity and eligibility criteria were inconsistent. We judged the risk of imprecision as moderate, due to the possible influence of sample size on proportion. Risk of publication bias evaluation identified symmetry and very low risk of bias in funnel plots (Supplementary Table 8; Supplementary Figures 10-11).

## Discussion

We systematically investigated the prevalence of radiological and functional consequences post-hospitalisation for viral pneumonitis, particularly for that caused by SARS-CoV-2. We observed the presence of inflammatory changes during hospitalisation in over 90% of CT scans, which reduced to 44% at a median follow up of 3 months. In contrast fibrotic changes were observed in a smaller percentage of CT scans (25-30%), though estimates remained more consistent between hospitalisation and follow-up, suggesting persistent fibrotic change. In analyses of lung function, restrictive impairment was estimated in 15%, whilst impaired gas transfer was observed in 39%. Heterogeneity in overall estimates were frequently substantial and therefore results should be interpreted with caution. We demonstrate that parenchymal lung damage by viral insult may be common and has the potential to explain COVID-19 related breathlessness in the months following hospitalisation.

We observed that overall estimates of radiological findings were consistent in sensitivity analysis restricted to the 3-6 months follow up time points. Least heterogeneity without adjustment was observed at four months, suggesting similar timeframes may be suitable for radiological follow-up. The presence of chronic respiratory disease diagnoses prior to hospitalisation may lead to overestimation in the prevalence of radiological changes. We addressed this by performing a sub analysis on studies that report radiological findings during hospitalisation, which supports interpretation of changes over time although we cannot confirm exclusion of individuals with underlying respiratory conditions.

A high proportion of people with inflammatory findings such as ground glass opacities and consolidation were observed at baseline following SARS-CoV-2, consistent with the radiological signs commonly described for viral pneumonitis.^118,119^ The difference in inflammatory changes reduced over the course of matched follow-up. Fibrotic changes were observed in a similar proportion of people during hospitalisation and at follow-up, suggesting a potential lack of resolution in the first year following infection. Meta regression indicated that, whilst not always significant, estimates of radiological sequelae reduced over time, particularly for inflammatory changes and more slowly for fibrotic changes. Radiological and functional sequelae have been described up to five years after Influenza infections,^15,114,120^ and up to fifteen years after SARS-CoV.^13,121,122^

In individuals with SARS-CoV-2, restrictive and gas transfer impairment were associated with infection severity,^40,42,43,63,70,123^ with similar findings reported in SARS-CoV,^88,94^ although not always statistically significant.^42,124^ We observe that the estimated prevalence of impaired gas transfer is greater than the prevalence of restrictive impairment following SARS-CoV-2 infection, with similar findings following other viral pneumonias. Meta-regression suggested that estimates of impaired gas transfer reduced over time, whilst the lower estimates of restrictive impairment did not change. Unresolved radiological changes and impaired lung function are important diagnostic tools for fibrotic interstitial lung disease, and prospective studies should accurately define the prevalence of post-COVID pulmonary fibrosis.^125^ (REF protocol when available)

Other systematic reviews have been published addressing radiological changes on CT and impairment to lung function in response to COVID-19, often limited to smaller numbers of studies, shorter follow-up, qualitative review alone, or lack of a preregistered protocol^126-129^ We included over 40 studies in quantitative synthesis of each radiological and physiological sequelae based on a preregistered protocol, including up to 12 months of follow-up, representing the largest systematic review and meta-analysis. High levels of heterogeneity are routinely reported in meta-analysis of proportions, so we perform sensitivity analysis, sub analysis and meta-regression to provide further reliable insights. We additionally model potential sources of heterogeneity in meta-regression, identifying timing of follow-up as an important characteristic to interpret estimates. A high risk of selection bias commonly contributed to variance in fibrotic estimates, whilst prospective design more commonly contributed to variance in inflammatory estimates, both of which highlight the impact of study inclusion criteria upon generalisability of systematic review findings. Unique to our protocol, we separately report estimates from Influenza and SARS-CoV studies, which suggest similar changes in response to non-COVID-19 viral pneumonitis.

There are limitations to this systematic review and meta-analysis. As our search strategy focused on follow-up tests, the number of included articles that reported baseline findings were limited, and no studies included CT findings prior to hospitalisation. Estimates of proportion are based on the number of tests performed, not patients infected, which would be affected by selection bias. Interpreting estimates requires caution as heterogeneity was frequently substantial and not completely attributable to the study-level features evaluated, consistent reasons for outlying study estimates were not identified. It is likely that variability in case mix demographic and severity of acute infection contributed to the heterogeneity between studies, which may be addressed by individual patient data approaches. All estimates represent individuals hospitalised with infection, which may not reflect prevalence in non-hospitalised cases. We defined radiological sequelae attributable to inflammatory and fibrotic responses, however these were not always reported specifically or exclusively and there are limitations to classifying radiological patterns. Ground-glass opacities are not exclusive to inflammation and could reflect retractile fibrosis during follow up. Approach to radiological classification only explained minor variance in fibrotic estimates, specific patterning likely contributes to residual heterogeneity. Internationally standardised approaches to reporting of post-COVID radiological change would support patient management and epidemiological study.

We have demonstrated the presence of substantial radiological and functional sequelae following viral pneumonias that may be consistent with post-viral interstitial lung disease. These parenchymal sequelae of viral infection could have a considerable impact given the large numbers of people discharged from hospital with COVID-19. Whilst the certainty of the presented estimates is low, they justify vigilant radiological and functional follow up of individuals hospitalised with viral pneumonia.

## Supporting information

PRISMA

## Data Availability

Data are publicly available in published manuscripts, and can be shared upon reasonable request

## Abbreviations

CT: computed tomography
DLCO: diffusing capacity for carbon monoxide
FEV1: forced expiratory volume in the first second
FVC: forced vital capacity
TLC: total lung capacity

## Contributions

IS had full access to all the data in the study and takes responsibility for the integrity of the data and the accuracy of the data analysis. LF, SM and IS have accessed and verified the underlying data. IS, GJ, AS and KR contributed to conceptualisation; IS, KR, JX, WC contributed to methodology; LF, SM, FK, WC contributed data curation; SM, FK, WC, JX contributed resource; LF, IS contributed formal analysis; IS, GJ contributed to supervision; GJ contributed funding acquisition; LF, IS contributed to writing – original draft. All authors contributed writing – review & editing. LF, SM, FK, WC, JX, KR, AS and IS report no competing interests relating to the manuscript, GJ reports NIHR BRC salaries, studentships, professorship (RP-2017-08-ST2-014). GJ also reports consulting fees and honoraria from Biogen, Galapagos, Galecto, GlaxoSmithKline, Heptares, MedImmuine, Pliant, PharmAkea, Bristol Myers Squibb, Veracyte, Boehringer Ingelheim, NordicBiosciences, Roche, Chiesi. The funder of the study (NIHR) had no role in study design, data collection, data analysis, data interpretation, or writing of the report. We acknowledge the following individuals for their correspondence and support in extracting individual study data: Dr Alexandra Kadl, Dr Ayham Daher, Dr Chris Ryerson, Dr David Arnold, Dr Edita Strumiliene, Dr Ernesto Crisafulli, Dr Fabio Anastasio, Dr Francesco Lombardi, Dr Gianluca Boari, Dr Giovanni Barisione, Dr Hosein Tabatabaei, Dr Jiang Yongpo, Dr Jose Alberto Fernandez Villar, Dr Li He, Dr Marco Marando, Dr Mattia Bellan, Dr Michiel de Graaf,Dr Mostafa Rasha, Dr Nick Maskell, Dr Rob van Gassel, Dr Susanne van Santen, Dr William Man.

