## Supplementary for "Post-viral parenchymal lung disease following COVID-19 and viral pneumonitis hospitalisation: A systematic review and meta-analysis"

Table of Content

Supplementary Table 1. Studies overview

**Centre: S: single centre; M: multicenter**

**Study design: P: prospective; R: retrospective**

**Method of diagnostic: ¶: PCR; ||: serology; *: other (reference to National guidelines)**

**Outcomes reported: †: radiological; ‡: restrictive; § reduction of DLCO; NA: not available**

**FU: Follow-up in months**

**Quantitative synthesis: n: not included; r: radiological outcome; f: functional outcome; b: both outcomes**

| **Author(s)** | **Year** | **Country** | **Centre** | **Study design** | **Sample size** | **Viral Agent** | **Method of diagnostic** | **Sex (males) N** | **Age reporting (years)** | **N ever smokers** | **FU months** | **Quantitative synthesis** |
| --- | --- | --- | --- | --- | --- | --- | --- | --- | --- | --- | --- | --- |
| Winterbauer et al. | 1977 | United States | S | P Case series | 11 | H3N2 | \|\| | 5 | mean+SD 62.09 (14.93) | NA | 1 | n |
| Antonio et al. | 2003 | China | S | R Cohort | 24 | SARS-CoV | * | 10 | mean+range 39 (23-70) | NA | 2 | r |
| Jin et al. | 2003 | China | S | R Cohort | 100 | SARS-CoV | * | 43 | mean+SD 36.7 (11.4) | NA | 2 | r |
| Liu, T., et al. | 2003 | China | M | R Cohort | 119 | SARS-CoV | * | 64 | mean+SD 34.1 (11.4) | NA | 1 | n |
| Peng et al. | 2003 | China | S | R Cohort | 89 | SARS-CoV | * | 40 | mean+SD 35.7 (11.1) | 23 | 2 | f |
| Chiang et al. | 2004 | Taiwan | S | P Cohort | 14 | SARS-CoV | ¶ | 3 | mean+SD 36.1 (13.9) | 0 | 8 | r |
| Hsu et al. | 2004 | Taiwan | S | R Cohort | 19 | SARS-CoV | ¶+ \|\| | 6 | mean+SD 42.5 (12.4) | 4 | 1 | r |
| Ng et al. | 2004 | China | S | P Cohort | 57 | SARS-CoV | ¶+ \|\| | 22 | mean+SD 38.1 (10.7) | 3 | 6 | f |
| Wong et al. | 2004 | China | S | R Cohort | 99 | SARS-CoV | \|\| | 41 | mean+SD 39.4 (12.8) | NA | 6 | r |
| Beijing Panel | 2005 | China | M | R Cohort | 456 | SARS-CoV | NA | 90 | mean+SD 33 (9) | NA | 6 | f |
| Chang et al. | 2005 | Taiwan | S | P Cohort | 40 | SARS-CoV | \|\| | 15 | mean+SD 42.8 (12.3) | 1 | 5 | r |
| Hui et al. | 2005 | China | S | P Cohort | 97 | SARS-CoV | \|\| | 39 | mean+SD 36.9 (9.5) | 3 | 12 | f |
| Ong et al. | 2005 | Singapore | S | P Cohort | 94 | SARS-CoV | \|\| | 24 | mean+SD 37 (12) | 7 | 12 | f |
| Xie et al. | 2005 | China | M | P Cohort | 383 | SARS-CoV | \|\| | 160 | mean+SD 38.2 (13.6) | NA | 12 | b |
| Zheng et al. | 2005 | China | S | R Cohort | 26 | SARS-CoV | * | 10 | mean+SD 36.2 (11.2) | NA | 18 | n |
| Chen et al. | 2006 | China | S | P RCT | 85 | SARS-CoV | * | 45 | mean+SD 37.2 (10.91) | NA | 2 | n |
| Li, T., et al. | 2006 | China | S | R Cohort | 59 | SARS-CoV | \|\| | 34 | mean+SD 47 (16) | 3 | 12 | n |
| Tansey et al. | 2007 | Canada | S | P Cohort | 117 | SARS-CoV | \|\| | 39 | median+IQR 42 (33-51) | 20 | 12 | n |
| Lu, et al. | 2010 | China | S | R Case series | 2 | H5N1 | ¶ | 1 | mean+SD 31.5 (7.78) |  | 12 | n |
| Bai et al. | 2011 | China | S | P Cohort | 65 | H1N1 | ¶ | 39 | median+IQR 41 (28-57) | 19 | 3 | r |
| Toufen et al. | 2011 | Brazil | S | P Cohort | 4 | H1N1 | ¶ | 4 | median+range 35.5 (25-54) | NA | 6 | n |
| Zarogoulidis et al. | 2011 | Greece | S | R Cohort | 44 | H1N1 | ¶ | 28 | mean+SD 36 (14.7) | 11 | 6 | n |
| Luyt et al. | 2012 | France | M | P Cohort | 37 | H1N1 | ¶ | 18 | median+IQR ECLA: 35.5 (30-39) - Non-ECLA: 42 (32.75-51.25) | 17 | 12 | r |
| Mineo et al | 2012 | Italy | S | R Case series | 20 | H1N1 | ¶ | 11 | mean+SD 43.5 (16.4) | NA | 999 | n |
| Quispe-Laime et al. | 2012 | Argentina | S | P Cohort | 7 | H1N1 | ¶ | 4 | mean+SD 37.43 (10.51) | 5 | 6 | n |
| Singh et al. | 2012 | India | S | R Case report | 1 | H1N1 | ¶ | 0 | specific value+NA 29 | NA | 12 | n |
| Edgeworth et al. | 2013 | Ireland | S | R Case series | 4 | H7N9 | ¶ | 3 | median+range 34.5 (31-56) | 1 | 19 | n |
| Dai et al. | 2014 | China | M | R Cohort | 10 | H1N1 | ¶ | 6 | mean+SD 53.6 (15.9) | NA | 3 | n |
| Liu, W., et al. | 2015 | China | S | R Cohort | 48 | H1N1 | * | 26 | mean+SD 29.5 (27-39.5) | NA | 12 | f |
| Wu, X. et al. | 2016 | China | M | R Case series | 11 | SARS-CoV | * | 3 | mean+range 38.6 (30-54) | 0 | 84 | r |
| Chen et al. | 2017 | China | S | P Cohort | 56 | H7N9 | * | 28 | mean+SD 54.48 (13.94) | 11 | 24 | b |
| Hsieh et al. | 2018 | Taiwan | S | P Cohort | 9 | H1N1 | ¶ | 9 | mean+SD 45.11 (5.48) | NA | 6 | n |
| Li, H., et al. | 2018 | China | M | P Cohort | 44 | H1N1-H7N9 | ¶ | 32 | median+IQR H7N9: 59 (48-73) - H7N1: 53 (40-62) | NA | 6 | r |
| Park, et al. | 2018 | South Korea | M | P Cohort | 73 | MERS-CoV | ¶ | 43 | median+range 51 (25-80) | 14 | 12 | f |
| Saha, et al. | 2018 | India | S | R Case series | 3 | H1N1 | ¶ ; \|\| | 3 | mean+SD 48 (9.85) | 0 | 12 | n |
| Anastasio et.al | 2021 | Italy | S | P Cohort | 222 | SARS-CoV-2 | ¶ | 127 | median+IQR 58(53-67) | 84 | 4 | f |
| Arnold et al. | 2020 | United Kingdom | S | P Cohort | 110 | SARS-CoV-2 | ¶ | 91 | median+IQR 60 (46-73) | NA | 3 | f |
| Barisione et al. | 2021 | Italy | S | P Cohort | 94 | SARS-CoV-2 | ¶ | 65 | mean+SD 61(12.1) | 52 | 1 | b |
| Bellan et al. | 2021 | Italy | S | P Cohort | 238 | SARS-CoV-2 | ¶ ; \|\| | 142 | median+IQR 61 (50-71) | 99 | 4 | f |
| Boari et al. | 2021 | Italy | S | P Cohort | 94 | SARS-CoV-2 | ¶ | 61 | mean+SD 66(11) | 15 | 4 | b |
| Bonnesen et al. | 2021 | Denmark | S | P Cohort | 12 | SARS-CoV-2 | * | 11 | median+IQR 62(57-67) | 9 | 3 | n |
| Cao et al. | 2021 | China | S | P Cohort | 81 | SARS-CoV-2 | ¶ | 47 | mean+SD 45(15) | 9 | 3 | b |
| Crisafulli et al | 2021 | Italy | S | P Cohort | 81 | SARS-CoV-2 | ¶ | 54 | mean+SD 66.5(11.2) | 35 | 4 | f |
| Daher et al. | 2020 | Germany | S | P Cohort | 33 | SARS-CoV-2 | ¶ | 22 | mean+SD 64 (3) | NA | 1.5 | f |
| de Graaf et al. | 2021 | Netherlands | S | P Cohort | 81 | SARS-CoV-2 | ¶ | 51 | mean+SD 61 (13) | 9 | 1.5 | n |
| Ekbom et al. | 2021 | Sweden | S | P Cohort | 60 | SARS-CoV-2 | ¶ | 43 | mean+range 59(27-82) | 14 | 4 | f |
| Finney at al. | 2021 | United Kingdom | S | P Cohort | 50 | SARS-CoV-2 | ¶ | 40 | median+IQR 54.5(44-59) | 14 | 1.5 | n |
| Frija-Masson et al. | 2021 | France | S | P Cohort | 137 | SARS-CoV-2 | ¶ | 69 | median+IQR 59(50-68) | 55 | 3 | b |
| Froidure et al | 2021 | Belgium | S | P Cohort | 134 | SARS-CoV-2 | ¶ | 79 | median+IQR 60(53-68) | 30 | 3 | b |
| Gianella et al. | 2021 | Switzerland | S | P Cohort | 39 | SARS-CoV-2 | ¶ | 30 | median+IQR 62.5 (51-71) | 15 | 3 | b |
| Gonzalez et al. | 2021 | Spain | S | P Cohort | 62 | SARS-CoV-2 | N/A | 46 | median+IQR 60 (48-65) | 34 | 3 | b |
| Gulati et al. | 2021 | USA | S | R Case series | 12 | SARS-CoV-2 | ¶ | 8 | mean+range 65.1(35-89) | 7 | 3 | n |
| Guler et al. | 2021 | Switzerland | M | P Cohort | 113 | SARS-CoV-2 | N/A | 67 | mean+SD 57.22 (12.11) | NA | 4 | r |
| Han et al. | 2021 | China | S | P Cohort | 114 | SARS-CoV-2 | ¶ | 80 | mean+SD 54 (12) | 16 | 6 | b |
| Huang C., et al. | 2021 | China | S | P Cohort | 1733 | SARS-CoV-2 | * | 897 | median+IQR 57 (47-65) | 146 | 6 | b |
| Huang, Y., et al. | 2020 | China | S | P Cross-Sectional | 57 | SARS-CoV-2 | ¶ | 26 | mean+SD 46.72 (13.78) | 9 | 1 | b |
| Labarca et al. | 2021 | Chile | M | P Cross-Sectional | 42 | SARS-CoV-2 | ¶ | 26 | mean+SD 48(10.75) | 22 | 4 | b |
| Lago et al. | 2021 | Brazil | S | R Case series | 4 | SARS-CoV-2 | N/A | 2 | median+SD 64(5.6) | 4 | 2 | n |
| Lerum et al. | 2021 | Norway | M | P Cohort | 103 | SARS-CoV-2 | ¶ | 54 | median+IQR 59(49-72) | 37 | 3 | b |
| Li, R., et al. | 2020 | China | S | R Cohort | 53 | SARS-CoV-2 | ¶ | 22 | mean+SD 50.2 (15.2) | NA | 8 | n |
| Li, X. et al. | 2021 | China | S | P Cohort | 289 | SARS-CoV-2 | ¶ |  | mean+SD 43.6(17.4) | N/A | 6 | b |
| Liang et al. | 2020 | China | S | P Cohort | 76 | SARS-CoV-2 | ¶ | 21 | mean+SR 41.3 (13.8) | 0 | 3 | f |
| Liu C., et al. | 2020 | China | S | R Cohort | 51 | SARS-CoV-2 | ¶ | 21 | mean+SR 46.6 (13.9) | 3 | 2 | r |
| Liu M., et al. | 2021 | China | S | P Cohort | 41 | SARS-CoV-2 | * | 22 | mean+SD 50(14) | N/A | 7 | r |
| Liu, D., et al. | 2020 | China | S | P Cohort | 149 | SARS-CoV-2 | ¶ | 82 | mean+IQR 43 (36-56) | NA | 1 | r |
| Liu, X., et al. | 2020 | China | S | R Cohort | 99 | SARS-CoV-2 | ¶ | 55 | means+SD 56.13 (20.7) | NA | 2 | n |
| Lombardi et al. | 2021 | Italy | S | P Cohort | 86 | SARS-CoV-2 | ¶ | 58 | mean+SD 58(13) | 52 | 1 | f |
| Lv et al. | 2020 | China | S | R Cohort | 137 | SARS-CoV-2 | ¶ | 71 | mean+SD 47 (13) | 6 | 0.5 | f |
| McGroder et al. | 2021 | USA | S | p Cohort | 76 | SARS-CoV-2 | ¶ | 45 | mean+SD 54(13.7) | 31 | 4 | r |
| Miwa et al. | 2021 | Japan | S | R Case series | 17 | SARS-CoV-2 | * | 14 | median+IQR 63(59-67) | 11 | 3 | n |
| Morin et al. | 2021 | France | S | P Cohort | 177 | SARS-CoV-2 | ¶ | 109 | mean+SD 56.9(13.2) | 40 | 4 | b |
| Myall et al. | 2021 | United Kingdom | S | P Cohort | 325 | SARS-CoV-2 | ¶ ; * | 25 | mean+SD 60.5 (10.7) | 21 | 1.5 | n |
| Noel-Savina et al. | 2021 | France | S | P Cohort | 72 | SARS-CoV-2 | ¶ | 55 | mean+SD 60.5(12.8) | 29 | 4 | b |
| Núñez-Fernández | 2021 | Spain | S | P Cohort | 225 | SARS-CoV-2 | ¶ | 119 | median+IQR 62(50-71) | 84 | 3 | f |
| Polese et al. | 2021 | Brazil | S | P Cohort | 41 | SARS-CoV-2 | ¶ | 30 | mean+SD 51(14) | 6 | 1 | f |
| Qin, W. et al. | 2021 | China | S | P Cohort | 81 | SARS-CoV-2 | ¶ | 34 | mean+SD 59(14) | N/A | 3 | b |
| Raman et al. | 2021 | United Kingdom | S | P Cohort | 58 | SARS-CoV-2 | ¶ | 34 | mean+SD 55.4(13.2) | 20 | 3 | f |
| Ramani et al. | 2021 | USA | S | P Case series | 28 | SARS-CoV-2 | ¶ | 17 | mean+SD 55.5 (11.9) | NA | 1.5 | f |
| Santus et al. | 2021 | Italy | S | P Cohort | 20 | SARS-CoV-2 | ¶ | 14 | mean+SD 58.3(15.5) | 4 | 1.5 | f |
| Schandl et al. | 2021 | Sweden | S | P Cohort | 113 | SARS-CoV-2 | ¶ | 86 | mean+SD 58(12.8) | 44 | 6 | f |
| Shah et al. | 2020 | Canada | S | P Cohort | 60 | SARS-CoV-2 | ¶ | 41 | median+IQR 67 (54-74) | 23 | 3 | b |
| Sibila et al. | 2021 | Spain | S | P Cohort | 172 | SARS-CoV-2 | ¶ | 98 | mean+SD 56.1(19.8) | 47 | 3 | f |
| Smet et al. | 2021 | Belgium | S | P Cross-Sectional | 220 | SARS-CoV-2 | NA | 135 | mean+SD 53 (13) | 52* | 1.5 | b |
| Strumiliene et al. | 2021 | Lithuania | S | P Cohort | 51 | SARS-CoV-2 | ¶ | 25 | mean+SD 56(11.72) | 4 | 2 | b |
| Tabatabaei et al. | 2020 | Iran | S | R Cohort | 52 | SARS-CoV-2 | * | 32 | mean+SD 50.17 (13.1) | 8 | 3 | r |
| van der Sar et al. | 2020 | Netherlands | S | P Cohort | 101 | SARS-CoV-2 | ¶ | 58 | mean+SD 66.4(12.6) | 56 | 1.5 | f |
| van Gassel et al. * | 2020 | Netherlands | S | P Cohort | 46* | SARS-CoV-2 | ¶ | 33 | median+ IQR 62 (55-68) | 21 | 7* | b |
| Wei et al. | 2020 | China | M | R Cohort | 59 | SARS-CoV-2 | * | 31 | mean+range 41 (25-70) | NA | 0.5 | r |
| Wu, Q. et al | 2021 | China | S | P Cohort | 54 | SARS-CoV-2 | ¶ | 32 | mean+SD 48(15.4) | N/A | 6 | b |
| Wu, X. et al. | 2021 | China | S | P Cohort | 83 | SARS-CoV-2 | ¶ | 47 | median+IQR 60(52-66) | 0 | 12 | b |
| Yasin et al. | 2021 | Egypt | S | R Cohort | 210 | SARS-CoV-2 | ¶ | 149 | mean+SD 53.85(24.8) | N/A | 2 | r |
| Yu et al. | 2020 | China | S | R Cohort | 32 | SARS-CoV-2 | ¶ | 22 | mean+SD 47.05 (17.85) | NA | 0.3 | r |
| Zhang, S. et al. | 2021 | China | S | R Cohort | 50 | SARS-CoV-2 | * | 19 | median+IQR 57(40-68) | 8 | 8 | b |
| Zhao et al. | 2020 | China | M | R Cohort | 55 | SARS-CoV-2 | ¶ | 32 | mean+SD 47.74 (15.49) | 4 | 3 | b |
| Zhong et al. | 2020 | China | S | R Cohort | 52 | SARS-CoV-2 | ¶ | 29 | mean+SD 45.46 (13.74) | NA | 1 | r |

Supplementary Table 2. Risk of bias assessment, observational studies.

Adapted from the CLARITY Group at McMaster University

| **Author(s)** | **Year** | **1. Can we be confident in the assessment of exposure?** | **2. Can we be confident that the outcome of interest was not present at start of study?** | **3. Did the study match the group(s) for all variables that are associated with the outcome of interest or did the statistical analysis adjust for these prognostic variables?** | **4. Can we be confident in the assessment of the presence or absence of prognostic factors?** | **5. Can we be confident in the assessment of outcome?** | **6. Was the follow up of cohorts adequate?** | **7. Were co-interventions similar between groups?** |
| --- | --- | --- | --- | --- | --- | --- | --- | --- |
| **SARS-CoV-2** |  |  |  |  |  |  |  |  |
| **Anastasio** | 2021 | V. Low | V. High | High | High | V. Low | High | N/A |
| **Arnold** | 2020 | V. Low | High | High | High | V. Low | Low | High |
| **Barisione** | 2021 | V. Low | Low | High | V. Low | V. Low | High | Low |
| **Bellan** | 2021 | V. Low | V. High | Low | High | V. Low | V. High | N/A |
| **Boari** | 2021 | V. Low | V. High | Low | Low | V. Low | V. High | N/A |
| **Bonnesen** | 2021 | Low | V. High | V. High | High | V. Low | High | N/A |
| **Cao** | 2021 | V. Low | V. High | High | High | V. Low | High | N/A |
| **Crisafulli** | 2021 | V. Low | V. High | Low | High | V. Low | V. Low | N/A |
| **Daher** | 2020 | V. Low | Low | V. High | Low | V. Low | V. Low | N/A |
| **de Graaf** | 2021 | V. Low | High | High | High | V. Low | Low | High |
| **Ekbom** | 2021 | V. Low | Low | Low | Low | V. Low | V. High | N/A |
| **Finney at al.** | 2021 | V. Low | V. High | Low | V. Low | V. Low | V. Low | N/A |
| **Frija-Masson** | 2021 | V. Low | V. Low | High | Low | V. Low | High | N/A |
| **Froidure** | 2021 | V. Low | V. Low | Low | Low | V. Low | Low | N/A |
| **Gianella** | 2021 | V. Low | Low | Low | Low | V. Low | V. Low | V. Low |
| **Gonzalez** | 2021 | Low | Low | V. Low | Low | V. Low | Low | N/A |
| **Gulati** | 2021 | V. Low | V. Low | V. High | High | V. Low | V. High | N/A |
| **Guler** | 2021 | V. High | Low | Low | Low | V. Low | High | High |
| **Han X** | 2021 | V. Low | High | Low | V. High | V. Low | High | Low |
| **Huang C** | 2021 | Low | High | Low | Low | V. Low | High | N/A |
| **Huang, Y** | 2020 | V. Low | Low | High | Low | V. Low | V. Low | Low |
| **Labarca** | 2021 | V. Low | High | V. Low | Low | V. Low | V. Low | Low |
| **Lago** | 2021 | V. High | Low | V. High | High | V. Low | V. Low | N/A |
| **Lerum** | 2021 | V. Low | V. High | Low | Low | V. Low | Low | N/A |
| **Li, R.** | 2020 | V. Low | High | V. High | V. High | V. Low | High | High |
| **Li, X.** | 2021 | V. Low | V. High | Low | High | High | High | High |
| **Liang, L** | 2020 | V. Low | High | Low | Low | V. Low | High | N/A |
| **Liu C** | 2020 | V. Low | V. High | V. High | High | V. Low | V. Low | N/A |
| **Liu D** | 2020 | V. Low | High | V. High | Low | V. Low | V. Low | N/A |
| **Liu M.** | 2021 | Low | V. Low | V. Low | V. Low | V. Low | V. Low | V. Low |
| **Liu X** | 2020 | V. Low | High | High | High | V. Low | High | High |
| **Lombardi** | 2021 | V. Low | High | Low | Low | V. Low | Low | N/A |
| **Lv** | 2020 | V. Low | V. High | N/A | V. High | V. Low | V. Low | N/A |
| **McGroder et al.** | 2021 | V. Low | V. Low | V. Low | V. Low | V. Low | High | V. Low |
| **Miwa** | 2021 | High | High | V. High | High | V. Low | High | N/A |
| **Morin** | 2021 | V. Low | High | V. Low | V. Low | V. Low | High | N/A |
| **Myall** | 2021 | Low | V. High | High | High | V. Low | V. High | N/A |
| **Noel-Savina** | 2021 | V. Low | Low | V. Low | Low | V. Low | Low | N/A |
| **Núñez-Fernández** | 2021 | V. Low | High | Low | V. Low | V. Low | V. Low | N/A |
| **Polese** | 2021 | V. Low | V. High | V. High | High | V. Low | High | N/A |
| **Qin** | 2021 | V. Low | Low | V. Low | V. Low | V. Low | V. High | V. Low |
| **Raman** | 2021 | V. Low | Low | V. Low | V. Low | V. Low | V. Low | High |
| **Ramani** | 2021 | Low | V. High | V. High | High | Low | High | N/A |
| **Santus** | 2021 | V. Low | V. Low | V. Low | V. Low | V. Low | High | N/A |
| **Schandl** | 2021 | V. Low | V. High | V. Low | V. Low | V. Low | V. High | Low |
| **Shah** | 2020 | V. Low | V. High | High | High | V. Low | High | N/A |
| **Sibila** | 2021 | V. Low | V. High | V. Low | High | V. Low | Low | N/A |
| **Smet** | 2021 | V. High | V. High | V. High | High | V. Low | High | Low |
| **Strumiliene** | 2021 | V. Low | Low | Low | Low | V. Low | Low | N/A |
| **Tabatabaei** | 2020 | Low | High | Low | Low | V. Low | V. Low | Low |
| **van der Sar - van der Brugge** | 2020 | V. Low | V. High | High | High | V. Low | Low | High |
| **van Gassel** | 2020 | V. Low | V. High | Low | Low | V. Low | High | N/A |
| **Wei J** | 2020 | Low | V. High | High | High | V. Low | High | High |
| **Wu, Q.** | 2021 | V. Low | V. Low | Low | High | V. Low | High | High |
| **Wu, X.** | 2021 | V. Low | Low | V. Low | V. Low | V. Low | High | N/A |
| **Yasin** | 2021 | V. Low | High | Low | High | V. Low | V. Low | N/A |
| **Yu M** | 2020 | V. Low | V. High | High | High | High | V. Low | High |
| **Zhang S.** | 2021 | V. Low | Low | Low | Low | V. Low | Low | N/A |
| **Zhao** | 2020 | V. Low | Low | Low | Low | V. Low | V. Low | Low |
| **Zhong L** | 2020 | Low | High | V. High | High | V. Low | V. High | High |
| **SARS-CoV** |  |  |  |  |  |  |  |  |
| **Antonio** | 2003 | Low | V. Low | High | High | Low | High | Low |
| **Beijing Respiratory Experts Pane** | 2005 | V. Low | V. Low | N/A | V. Low | V. Low | Low | N/A |
| **Chang** | 2005 | V. Low | Low | High | Low | Low | High | V. Low |
| **Chiang** | 2004 | V. Low | High | N/A | Low | Low | Low | N/A |
| **Hsu** | 2004 | V. Low | Low | High | High | V. Low | High | V. Low |
| **Hui D. S.** | 2005 | V. Low | Low | Low | Low | Low | High | Low |
| **Jin, Z. Y.** | 2003 | V. Low | V. Low | N/A | Low | Low | Low | N/A |
| **Li, T. S.** | 2006 | V. Low | High | Low | High | Low | Low | N/A |
| **Liu, T.** | 2003 | V. Low | V. Low | High | Low | V. Low | High | N/A |
| **Ng** | 2004 | V. Low | High | Low | High | V. Low | Low | V. Low |
| **Ong** | 2005 | Low | Low | V. Low | V. Low | V. Low | High | N/A |
| **Peng** | 2003 | V. Low | V. Low | Low | Low | High | V. Low | N/A |
| **Tansey** | 2007 | V. Low | V. Low | High | Low | V. Low | High | N/A |
| **Wong** | 2004 | V. Low | High | V. Low | High | V. Low | V. Low | N/A |
| **Wu X.** | 2016 | Low | High | N/A | High | V. Low | V. Low | N/A |
| **Xie, L.** | 2005 | V. Low | Low | Low | High | Low | Low | Low |
| **Zheng** | 2005 | V. Low | V. Low | N/A | Low | Low | V. High | N/A |
| **MERS-CoV** |  |  |  |  |  |  |  |  |
| **Park** | 2018 | V. Low | Low | N/A | High | V. Low | High | High |
| **Influenza** |  |  |  |  |  |  |  |  |
| **Bai, L.** | 2011 | Low | High | Low | Low | Low | V. High | High |
| **Chen, J.** | 2017 | High | High | Low | High | Low | Low | Low |
| **Dai, J.** | 2014 | Low | V. Low | N/A | V. High | V. Low | High | N/A |
| **Edgeworth, D.** | 2013 | Low | High | N/A | V. High | V. Low | N/A | N/A |
| **Hsieh** | 2018 | V. Low | Low | V. High | V. High | V. Low | V. Low | N/A |
| **Li, H.** | 2018 | V. Low | V. Low | Low | High | V. Low | V. Low | Low |
| **Liu, W.** | 2015 | High | Low | N/A | V. High | High | High | N/A |
| **Lu PX** | 2010 | V. Low | High | N/A | High | V. Low | V. Low | N/A |
| **Luyt** | 2012 | V. Low | V. Low | High | High | Low | V. Low | V. Low |
| **Mineo** | 2012 | V. Low | High | V. High | Low | V. Low | Low | N/A |
| **Quispe-Laime** | 2012 | V. Low | Low | N/A | V. High | Low | V. Low | N/A |
| **Saha** | 2018 | V. Low | High | N/A | V. High | Low | V. Low | Low |
| **Singh, V.** | 2012 | V. Low | High | N/A | V. High | Low | N/A | N/A |
| **Toufen, C.** | 2011 | V. Low | Low | N/A | V. High | Low | V. Low | N/A |
| **Winterbauer** | 1977 | V. Low | High | V. High | V. High | V. Low | Low | N/A |
| **Zarogoulidis** | 2011 | V. Low | Low | N/A | Low | Low | V. Low | N/A |

**Answer format:**

Very high. Definitely High risk of bias: There is direct evidence of high risk of bias practices.

High. Probably High risk of bias: There is indirect evidence of high risk of bias practices OR there is insufficient information provided about relevant risk of bias practices.

Low. Probably Low risk of bias: There is indirect evidence of low risk of bias practices OR it is deemed by the risk of bias evaluator that deviations from low risk of bias practices for these criteria during the study would not appreciably bias results, including consideration of direction and magnitude of bias

Very low. Definitely Low risk of bias: There is direct evidence of low risk of bias practices

Supplementary Table 3. Risk of bias assessment, randomized control trial.

From the CLARITY Group at McMaster University

| **Author(s)** | **Year** | **1. Was the allocation sequence adequately generated?** | **2. Was the allocation adequately concealed?** | **3. Blinding: Was knowledge of the allocated interventions adequately prevented?** | **4. Was loss to follow-up (missing outcome data) infrequent?** | **5. Are reports of the study free of selective outcome reporting?** | **6. Was the study apparently free of other problems that could put it at a risk of bias?** |
| --- | --- | --- | --- | --- | --- | --- | --- |
| **Chen, J., et al.** | 2006 | V. Low | High | High | V. Low | V. Low | Low |

**Answer format:**

Very high. Definitely High risk of bias: There is direct evidence of high risk of bias practices.

High. Probably High risk of bias: There is indirect evidence of high risk of bias practices OR there is insufficient information provided about relevant risk of bias practices.

Low. Probably Low risk of bias: There is indirect evidence of low risk of bias practices OR it is deemed by the risk of bias evaluator that deviations from low risk of bias practices for these criteria during the study would not appreciably bias results, including consideration of direction and magnitude of bias

Very low. Definitely Low risk of bias: There is direct evidence of low risk of bias practices

Supplementary Table 4. Meta regression in estimates of inflammatory changes

| All follow up (n=31) | |  |  |  |  |  |
| --- | --- | --- | --- | --- | --- | --- |
| **Covariate** | **Unit** | **Coefficient** | **95%CI** | **p value** | **Residual I^2^** | **R^2^** |
| **Time** | Months | -0.036 | -0.68; -0.004 | 0.029 | 73.1% | 14.7% |
| **Severity** | Mild | *Ref* |  |  |  |  |
|  | Moderate | -0.068 | -0.551; 0.415 | 0.782 |  |  |
|  | Severe | -0.061 | -0.570; 0.449 | 0.815 | 78.0% | 0.0% |
| **Design** | Retrospective | *Ref* |  |  |  |  |
|  | Prospective | 0.044 | -0.172; 0.261 | 0.688 | 77.2% | 0.0% |
| **Selection bias** | Weak | *Ref* |  |  |  |  |
|  | Strong | 0.009 | -0.157; 0.174 | 0.919 | 76.8% | 0.0% |
| 3-6 month follow up (n=19) | |  |  |  |  |  |
| **Covariate** | **Unit** | **Coefficient** | **95%CI** | **p value** | **Residual I^2^** | **R^2^** |
| **Time** | Months | -0.066 | -0.152; 0.020 | 0.133 | 71.0% | 9.3% |
| **Severity** | Mild | *Ref* |  |  |  |  |
|  | Moderate | -0.085 | -0.312; 0.142 | 0.461 | 75.3% | 0.0% |
|  | Severe | - |  |  |  |  |
| **Design** | Retrospective | *Ref* |  |  |  |  |
|  | Prospective | 0.271 | -0.050; 0.591 | 0.098 | 72.4% | 11.7% |
| **Selection bias** | Weak | *Ref* |  |  |  |  |
|  | Strong | -0.131 | -0.327; 0.066 | 0.192 | 73.7% | 2.6% |
| **Radiological Classification** | Study | *Ref* |  |  |  |  |
|  | Review | -0.071 | -0.276; 0.133 | 0.496 | 75.30% | 0.00% |
| matched follow up (n=12) | |  |  |  |  |  |
| **Covariate** | **Unit** | **Coefficient** | **95%CI** | **p value** | **Residual I^2^** | **R^2^** |
| **Time** | Months | -0.03 | -0.074; 0.014 | 0.185 | 55.7% | 0.0% |
| **Severity** | Mild | *Ref* |  |  |  |  |
|  | Moderate | -0.144 | -0.518; 0.229 | 0.448 |  |  |
|  | Severe | -0.085 | -0.085; 0.213 | 0.692 | 61.8% | 0.0% |
| **Design** | Retrospective | *Ref* |  |  |  |  |
|  | Prospective | -0.124 | -0.331; 0.082 | 0.237 | 55.9% | 5.3% |
| **Selection bias** | Weak | *Ref* |  |  |  |  |
|  | Strong | -0.025 | -0.218; 0.168 | 0.799 | 59.1% | 0.0% |
| **Radiological Classification** | Study | *Ref* |  |  |  |  |
|  | Review | -0.020 | -0.213; 0.174 | 0.843 | 59.6% | 0.0% |

Supplementary Table 5. Meta regression in estimates of fibrotic changes

| All follow up (n=33) | |  |  |  |  |  |
| --- | --- | --- | --- | --- | --- | --- |
| **Covariate** | **Unit** | **Coefficient** | **95%CI** | **p value** | **Residual I^2^** | **R^2^** |
| **Time** | Months | -0.021 | -0.051; 0.009 | 0.176 | 70.3% | 4.9% |
| **Severity** | Mild | *Ref* |  |  |  |  |
|  | Moderate | -0.097 | -0.541; 0.346 | 0.666 |  |  |
|  | Severe | -0.033 | -0.500; 0.435 | 0.891 | 73.3% | 0.0% |
| **Design** | Retrospective | *Ref* |  |  |  |  |
|  | Prospective | 0.079 | -0.120; 0.278 | 0.436 | 72.5% | 0.0% |
| **Selection bias** | Weak | *Ref* |  |  |  |  |
|  | Strong | -0.114 | -0.255; 0.027 | 0.113 | 69.8% | 6.5% |
| 3-6 month follow up (n=20) | |  |  |  |  |  |
| **Covariate** | **Unit** | **Coefficient** | **95%CI** | **p value** | **Residual I^2^** | **R^2^** |
| **Time** | Months | -0.082 | -0.158; -0.006 | 0.035 | 63.7% | 21.0% |
| **Severity** | Mild | *Ref* |  |  |  |  |
|  | Moderate | -0.113 | -0.319; 0.092 | 0.278 | 70.4% | 0.0% |
|  | Severe | - |  |  |  |  |
| **Design** | Retrospective | *Ref* |  |  |  |  |
|  | Prospective | 0.111 | -0.207; 0.429 | 0.493 | 72.1% | 0.0% |
| **Selection bias** | Weak | *Ref* |  |  |  |  |
|  | Strong | -0.176 | -0.343; -0.009 | 0.039 | 65.1% | 21.1% |
| **Radiological Classification** | Study | *Ref* |  |  |  |  |
|  | Review | -0.125 | -0.302; 0.053 | 0.169 | 69.3% | 2.9% |
| matched follow up (n=12) | |  |  |  |  |  |
| **Covariate** | **Unit** | **Coefficient** | **95%CI** | **p value** | **Residual I^2^** | **R^2^** |
| **Time** | Months | -0.026 | -0.072; 0.020 | 0.266 | 58.3% | 5.4% |
| **Severity** | Mild | *Ref* |  |  |  |  |
|  | Moderate | -0.116 | -0.516; 0.285 | 0.572 |  |  |
|  | Severe | -0.172 | -0.623; 0.279 | 0.454 | 67.6% | 0.0% |
| **Design** | Retrospective | *Ref* |  |  |  |  |
|  | Prospective | -0.010 | -0.239; 0.219 | 0.933 | 65.8% | 0.0% |
| **Selection bias** | Weak | *Ref* |  |  |  |  |
|  | Strong | -0.134 | -0.325; 0.057 | 0.17 | 58.2% | 8.3% |
| **Radiological Classification** | Study | *Ref* |  |  |  |  |
|  | Review | -0.108 | -0.323; -0.106 | 0.323 | 64.0% | 0.0% |

Supplementary Table 6. Meta regression in estimates of impaired gas transfer

| All follow up (n=35) | |  |  |  |  |  |
| --- | --- | --- | --- | --- | --- | --- |
| **Covariate** | **Unit** | **Coefficient** | **95%CI** | **p value** | **Residual I^2^** | **R^2^** |
| **Time** | Months | -0.018 | -0.046; 0.010 | 0.207 | 63.2% | 1.8% |
| **Severity** | Mild | *Ref* |  |  |  |  |
|  | Moderate | -0.205 | -0.649; 0.231 | 0.358 |  |  |
|  | Severe | -0.152 | -0.604; 0.299 | 0.509 | 64.9% | 0.0% |
| **Design** | Retrospective | *Ref* |  |  |  |  |
|  | Prospective | 0.114 | -0.093; 0.321 | 0.280 | 62.7% | 0.0% |
| **Selection bias** | Weak | *Ref* |  |  |  |  |
|  | Strong | 0.017 | -0.101; 0.134 | 0.781 | 64.5% | 0.0% |
| 3-6 month follow up (n=24) | |  |  |  |  |  |
| **Covariate** | **Unit** | **Coefficient** | **95%CI** | **p value** | **Residual I^2^** | **R^2^** |
| **Time** | Months | -0.045 | -0.104; 0.014 | 0.136 | 59.4% | 6.7% |
| **Severity** | Mild | *Ref* |  |  |  |  |
|  | Moderate | -0.095 | -0.256; 0.066 | 0.248 | 62.0% | 0.2% |
|  | Severe | - |  |  |  |  |
| **Design** | Retrospective | *Ref* |  |  |  |  |
|  | Prospective | 0.112 | -0.088; 0.312 | 0.272 | 60.8% | 0.0% |
| **Selection bias** | Weak | *Ref* |  |  |  |  |
|  | Strong | 0.048 | -0.084; 0.180 | 0.474 | 62.4% | 0.0% |

Supplementary Table 7. Meta regression in estimates of restrictive impairment

| All follow up (n=33) | |  |  |  |  |  |
| --- | --- | --- | --- | --- | --- | --- |
| **Covariate** | **Unit** | **Coefficient** | **95%CI** | **p value** | **Residual I^2^** | **R^2^** |
| **Time** | Months | -0.002 | -0.031; 0.026 | 0.864 | 60.1% | 0.0% |
| **Severity** | Mild | *Ref* |  |  |  |  |
|  | Moderate | 0.018 | -0.406; 0.442 | 0.935 |  |  |
|  | Severe | 0.123 | -0.319; 0.564 | 0.586 | 60.5% | 0.0% |
| **Design** | Retrospective | *Ref* |  |  |  |  |
|  | Prospective | 0.086 | -0.113; 0.285 | 0.397 | 58.9% | 0.0% |
| **Selection bias** | Weak | *Ref* |  |  |  |  |
|  | Strong | -0.021 | -0.140; 0.098 | 0.728 | 60.1% | 0.0% |
| 3-6 month follow up (n=22) | |  |  |  |  |  |
| **Covariate** | **Unit** | **Coefficient** | **95%CI** | **p value** | **Residual I^2^** | **R^2^** |
| **Time** | Months | 0.001 | -0.041; 0.042 | 0.981 | 16.7% | 0.0% |
| **Severity** | Mild | *Ref* |  |  |  |  |
|  | Moderate | -0.108 | -0.228; 0.011 | 0.075 | 7.8% | 35.5% |
|  | Severe | - |  |  |  |  |
| **Design** | Retrospective | *Ref* |  |  |  |  |
|  | Prospective | 0.032 | -0.097; 0.160 | 0.63 | 18.2% | 0.0% |
| **Selection bias** | Weak | *Ref* |  |  |  |  |
|  | Strong | 0.052 | -0.036; 0.141 | 0.248 | 12.1% | 0.0% |

### Supplementary Table 8. GRADE approach to rate confidence in the estimates

| **No of studies** | | **Design** | **Risk of bias** | **Inconsistency** | **Indirectness** | **Imprecision** | **Publication bias** | **Certainty**  **(overall score)^[1]^** |
| --- | --- | --- | --- | --- | --- | --- | --- | --- |
| **a** | **Outcome:** Radiological sequelae - Fibrotic | | | | | | | |
| 45 | | Observational studies | Low to Moderate | Moderate | No serious indirectness | Moderate imprecision | Very low | 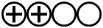  **Low** |
| **b** | **Outcome:** Radiological sequelae – Inflammatory | | | | | | | |
| 42 | | Observational studies | Low to Moderate | Moderate | No serious indirectness | Moderate imprecision | Very low | 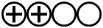  **Low** |
| **c** | **Outcome:** Functional sequelae – Restrictive impairment | | | | | | | |
| 39 | | Observational studies | Low to Moderate | Moderate | No serious indirectness | Moderate imprecision | Very low | 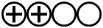  **Low** |
| **d** | **Outcome:** Functional sequelae – DLCO reduction | | | | | | | |
| 44 | | Observational studies | Low to Moderate | Moderate | No serious indirectness | Moderate imprecision | Very low | 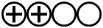  **Low** |

**Grading of Recommendations, Assessment, Development and Evaluations**

Risk of bias: Included studies with low to moderate risk of bias. Possible confounding factors were not extensively assessed.

Inconsistency: Moderate inconsistency with substantial heterogeneity that was largely explained by timing of follow up.

Indirectness: No serious indirectness. All study subjects had confirmed viral pneumonia, although severity and eligibility criteria were inconsistent.

Imprecision: Possible effect of sample size and selection.

Publication bias: Very low, funnel plots largely symmetrical with most studies within 95%CI threshold.

Supplementary Figure 1. MEDLINE search strategy (OVID). Last carried out on 1^st^ March 2021

1. viral pneumonia.mp. or Pneumonia, Viral/

2. Coronaviridae Infections/ or Coronavirus Infections/ or Infectious bronchitis virus/ or Coronaviridae/ or coronav*.mp. or Middle East Respiratory Syndrome Coronavirus/ or Coronavirus/

3. Betacoronavirus/ or Betacoronavirus 1/ or betacoronav*.mp.

4. severe acute respiratory syndrome.mp. or Severe Acute Respiratory Syndrome/

5. SARS Virus/ or sars.mp.

6. sars-cov-2.mp.

7. covid19.mp.

8. covid*.mp.

9. mers.mp. or Middle East Respiratory Syndrome Coronavirus/

10. Influenza A Virus, H7N9 Subtype/ or h7n9.mp. or Influenza A Virus, H5N1 Subtype/ or Influenza A virus/ or Influenza A Virus, H3N2 Subtype/ or Influenza A Virus, H1N2 Subtype/ or influenza*.mp. or Influenza A Virus, H1N1 Subtype/ or Influenza, Human/

11. Hospitalization/ or hospital*.mp.

12. Hypoxia/ or Oxygen Inhalation Therapy/ or oxygen therapy.mp.

13. Respiration, Artificial/ or invasive mechanical ventilation.mp. or Positive-Pressure Respiration/ or Respiratory Insufficiency/

14. Ventilation/ or Intermittent Positive-Pressure Ventilation/ or Noninvasive Ventilation/ or ventilation.mp. or Pulmonary Ventilation/

15. Noninvasive Ventilation/ or niv.mp.

16. Continuous Positive Airway Pressure/ or cpap.mp. or Positive-Pressure Respiration/

17. steroid*.mp.

18. Steroids/ or steroids*.mp.

19. Glucocorticoids/ or glucorticoids.mp.

20. antiviral.mp. or Antiviral Agents/

21. spirometry.mp. or Spirometry/

22. forced vital capacity.mp. or Vital Capacity/

23. fvc.mp.

24. forced expiratory volume.mp. or Forced Expiratory Volume/

25. fev1.mp.

26. total lung capacity.mp. or Total Lung Capacity/

27. tlc.mp.

28. dlco.mp. or Pulmonary Diffusing Capacity/

29. tlco.mp.

30. pulmonary function test.mp. or Respiratory Function Tests/

31. lung function*.mp.

32. Tomography, X-Ray Computed/ or ct.mp.

33. computed tomography.mp.

34. computer assisted tomography.mp.

35. fibrosis.mp. or Fibrosis/ or Pulmonary Fibrosis/

36. Bronchiolitis Obliterans/ or lung fibro*.mp.

37. hrct.mp.

38. high resolution computer tomography.mp. or Lung Diseases, Interstitial/

39. reticulation.mp.

40. fibrosing alveolitis.mp. or Pulmonary Fibrosis/

41. traction bronchiectasis.mp.

42. ground glass*.mp.

43. ggo.mp.

44. honeycombing.mp.

45. septal thickening.mp.

46. lung distortion.mp.

47. fibrotic disease.mp.

48. restrictive impairment.mp.

49. restrictive test.mp.

50. Cryptogenic Organizing Pneumonia/ or cryptogenic organising pneumonia.mp.

51. bronchiolitis obliterans organizing pneumonia.mp. or Cryptogenic Organizing Pneumonia/

52. 1 or 2 or 3 or 4 or 5 or 6 or 7 or 8 or 9 or 10

53. 11 or 12 or 13 or 14 or 15 or 16 or 17 or 18 or 19 or 20

54. 21 or 22 or 23 or 24 or 25 or 26 or 27 or 28 or 29 or 30 or 31 or 32 or 33 or 34 or 35 or 36 or 37 or 38 or 39 or 40 or 41 or 42 or 43 or 44 or 45 or 46 or 47 or 48 or 49 or 50 or 51

55. 52 and 53 and 54

56. limit 55 to (humans and "all adult (19 plus years)")

Supplementary Figure 2a. Risk of bias assessment, observational studies

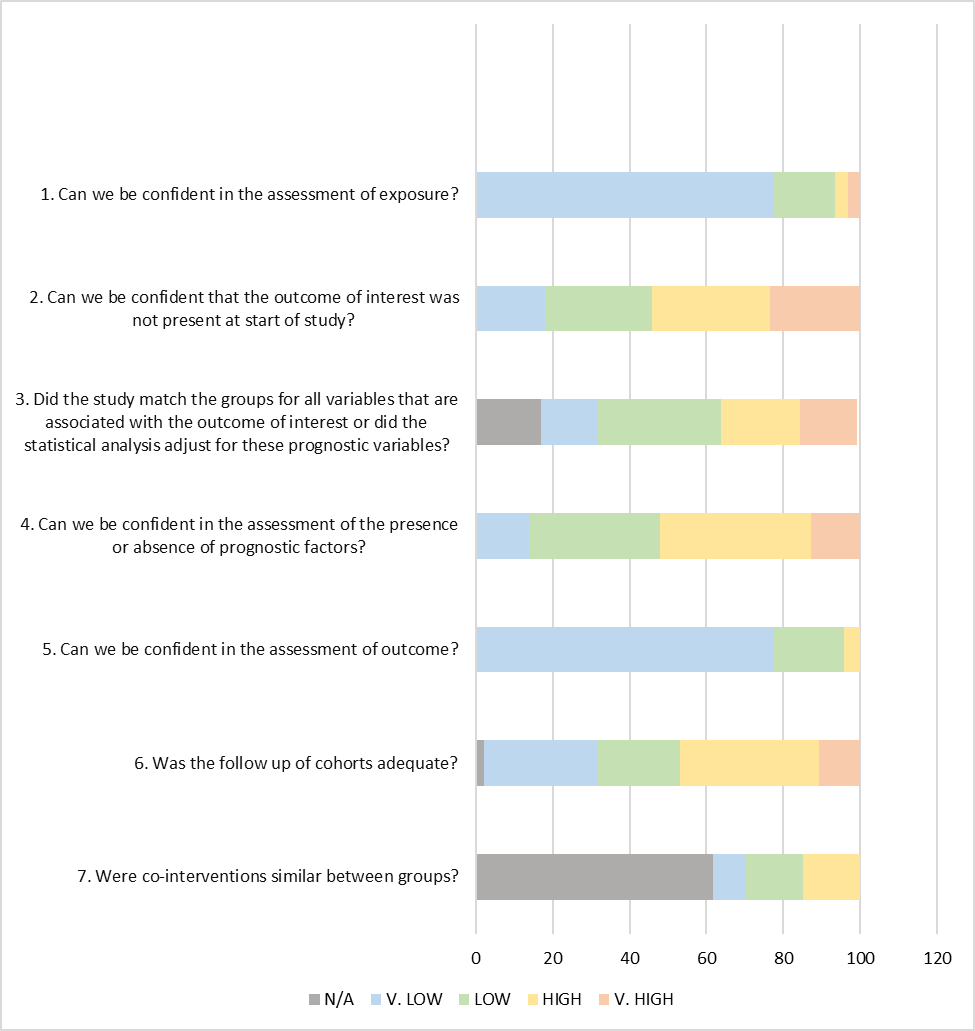

**Answer format:**

**Very high.** Definitely High risk of bias: There is direct evidence of high risk of bias practices.

**High.** Probably High risk of bias: There is indirect evidence of high risk of bias practices OR there is insufficient information provided about relevant risk of bias practices.

**Low**. Probably Low risk of bias: There is indirect evidence of low risk of bias practices OR it is deemed by the risk of bias evaluator that deviations from low risk of bias practices for these criteria during the study would not appreciably bias results, including consideration of direction and magnitude of bias

**Very low**. Definitely Low risk of bias: There is direct evidence of low risk of bias practices

### **Supplementary Figure 2b. Risk of bias assessment, observational studies**. Only SARS-CoV-2

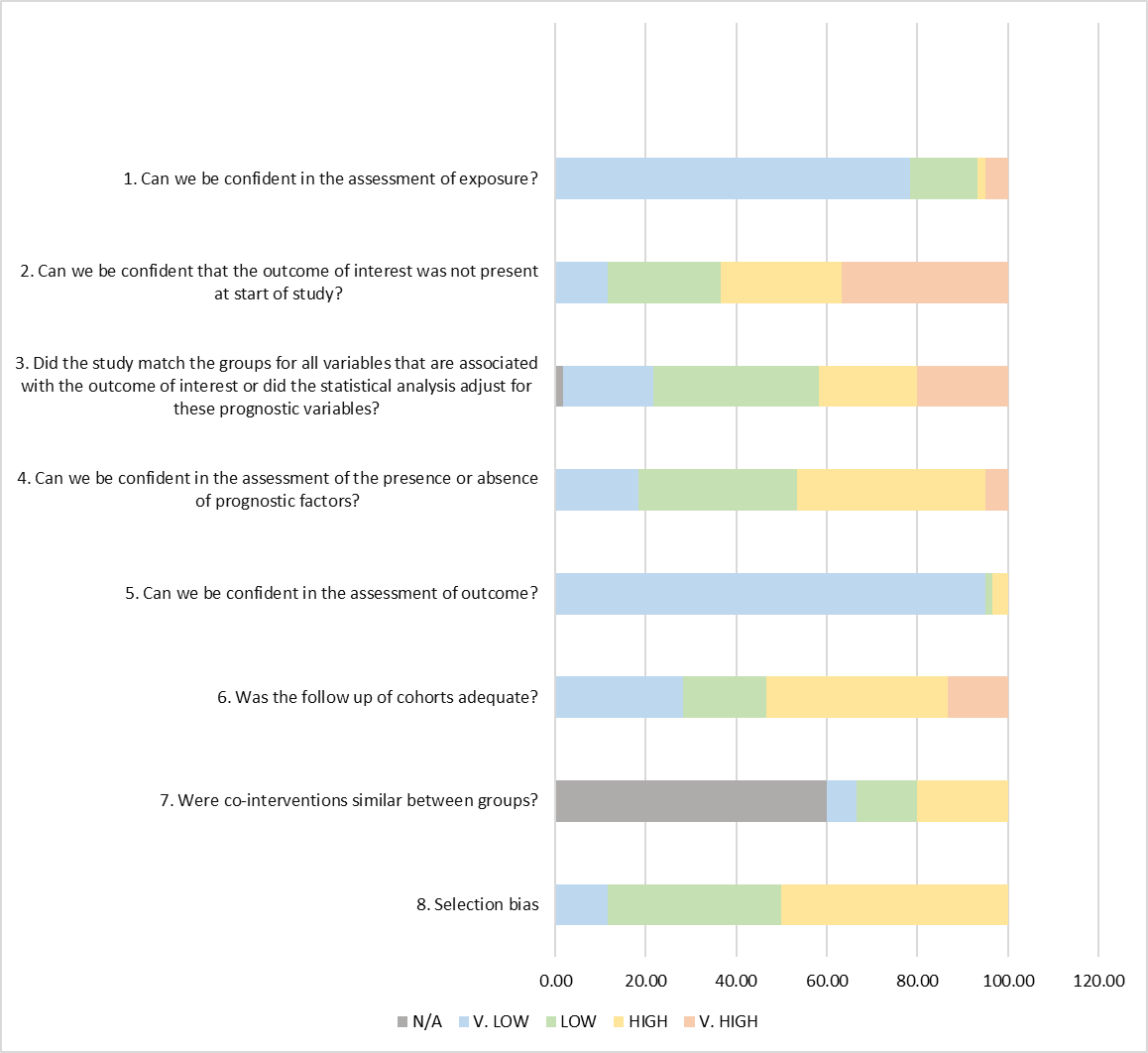

**Answer format:**

**Very high.** Definitely High risk of bias: There is direct evidence of high risk of bias practices.

**High.** Probably High risk of bias: There is indirect evidence of high risk of bias practices OR there is insufficient information provided about relevant risk of bias practices.

**Low**. Probably Low risk of bias: There is indirect evidence of low risk of bias practices OR it is deemed by the risk of bias evaluator that deviations from low risk of bias practices for these criteria during the study would not appreciably bias results, including consideration of direction and magnitude of bias

**Very low**. Definitely Low risk of bias: There is direct evidence of low risk of bias practices

### Supplementary Figure 3. Radiological findings at 3-6 months restricted follow-up: sensitivity analysis

Estimates are reported as proportion of number of CT scans showing the outcome of interest (n) on the total number of exams performed (N) and 95% confidence interval.

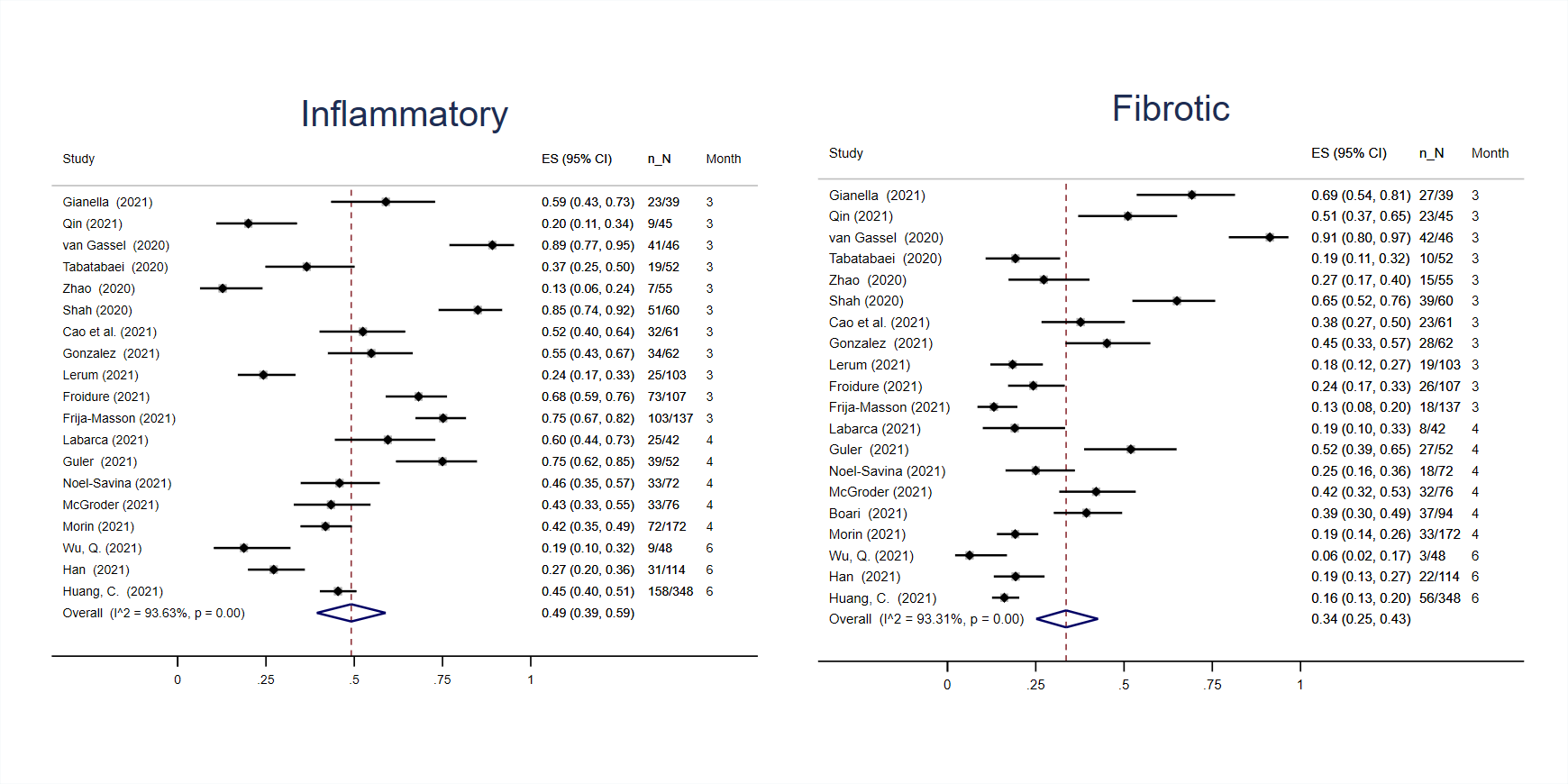

### Supplementary Figure 4. Radiological findings at 3-6 months restricted follow-up: stratified

Estimates are reported as proportion of number of CT scans showing the outcome of interest (n) on the total number of exams performed (N) and 95% confidence interval.

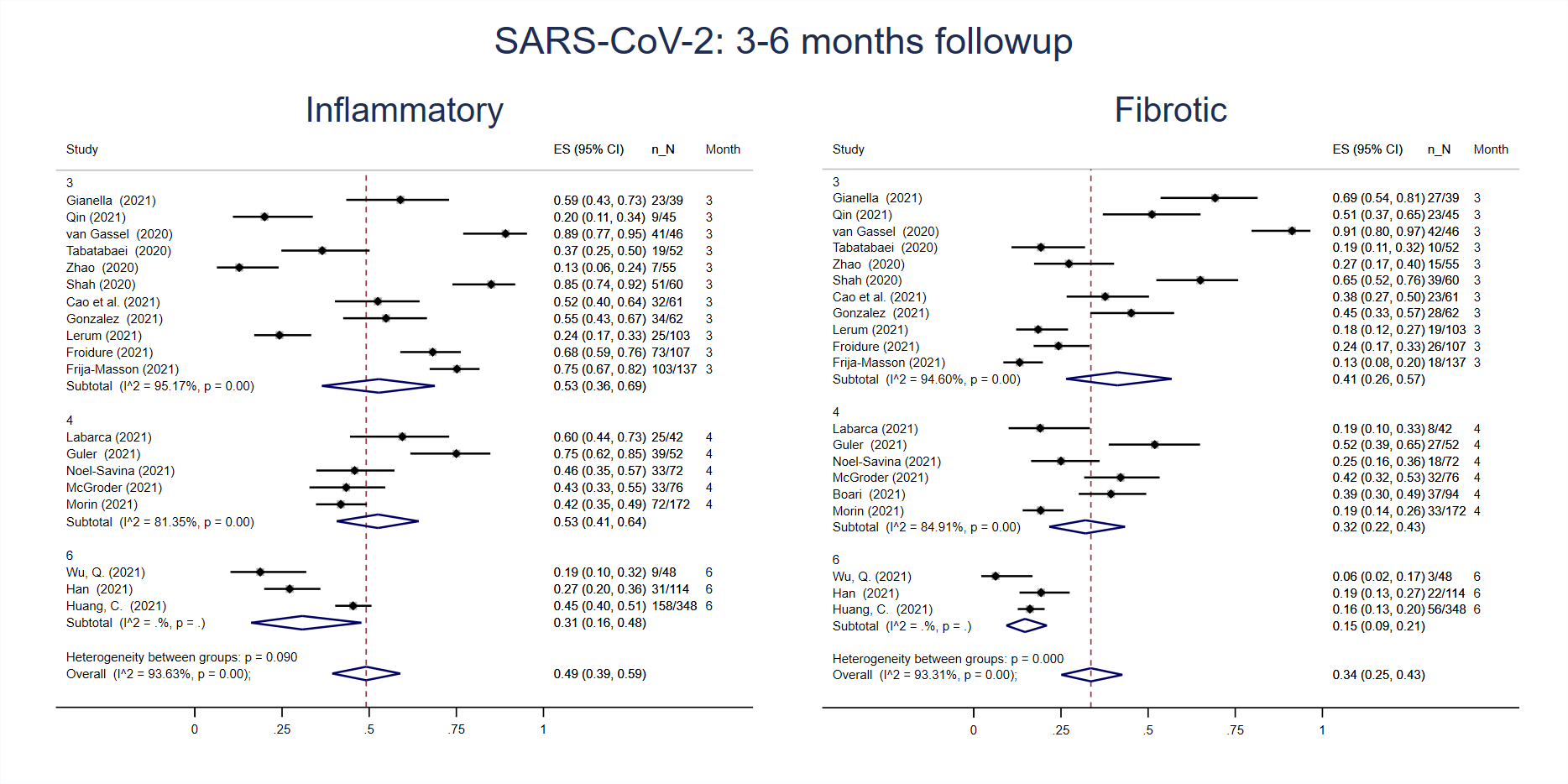

### Supplementary Figure 5. Radiological findings at follow-up in Influenza and SARS-CoV studies

Estimates are reported as proportion of number of CT scans showing the outcome of interest (n) on the total number of exams performed (N) and 95% confidence interval.

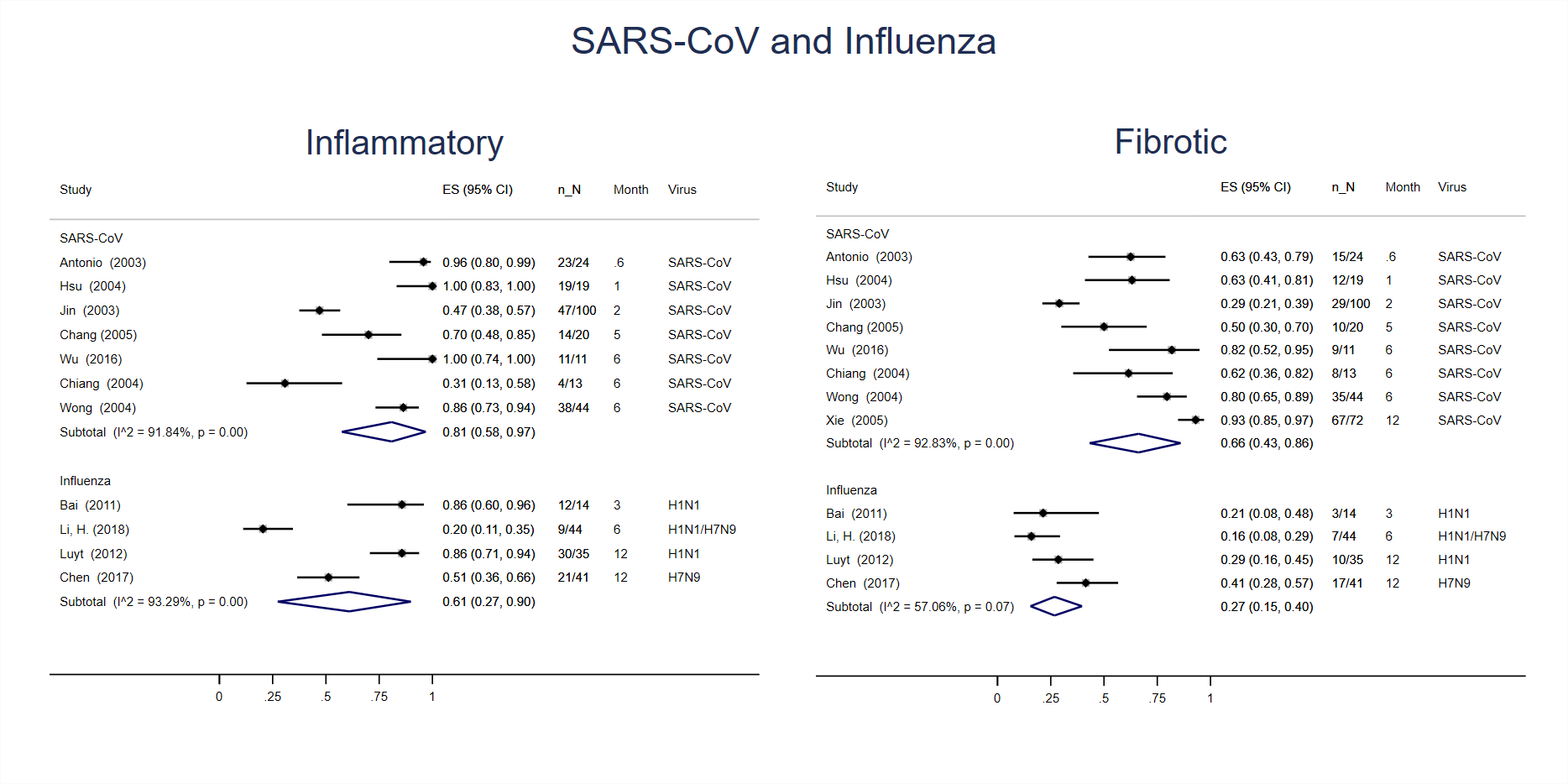

### Supplementary Figure 6. Lung function findings at 3-6 month restricted follow-up: sensitivity analysis

Estimates are reported as proportion of number of lung function tests showing the outcome of interest (n) on the total number of exams performed (N) and 95% confidence interval.

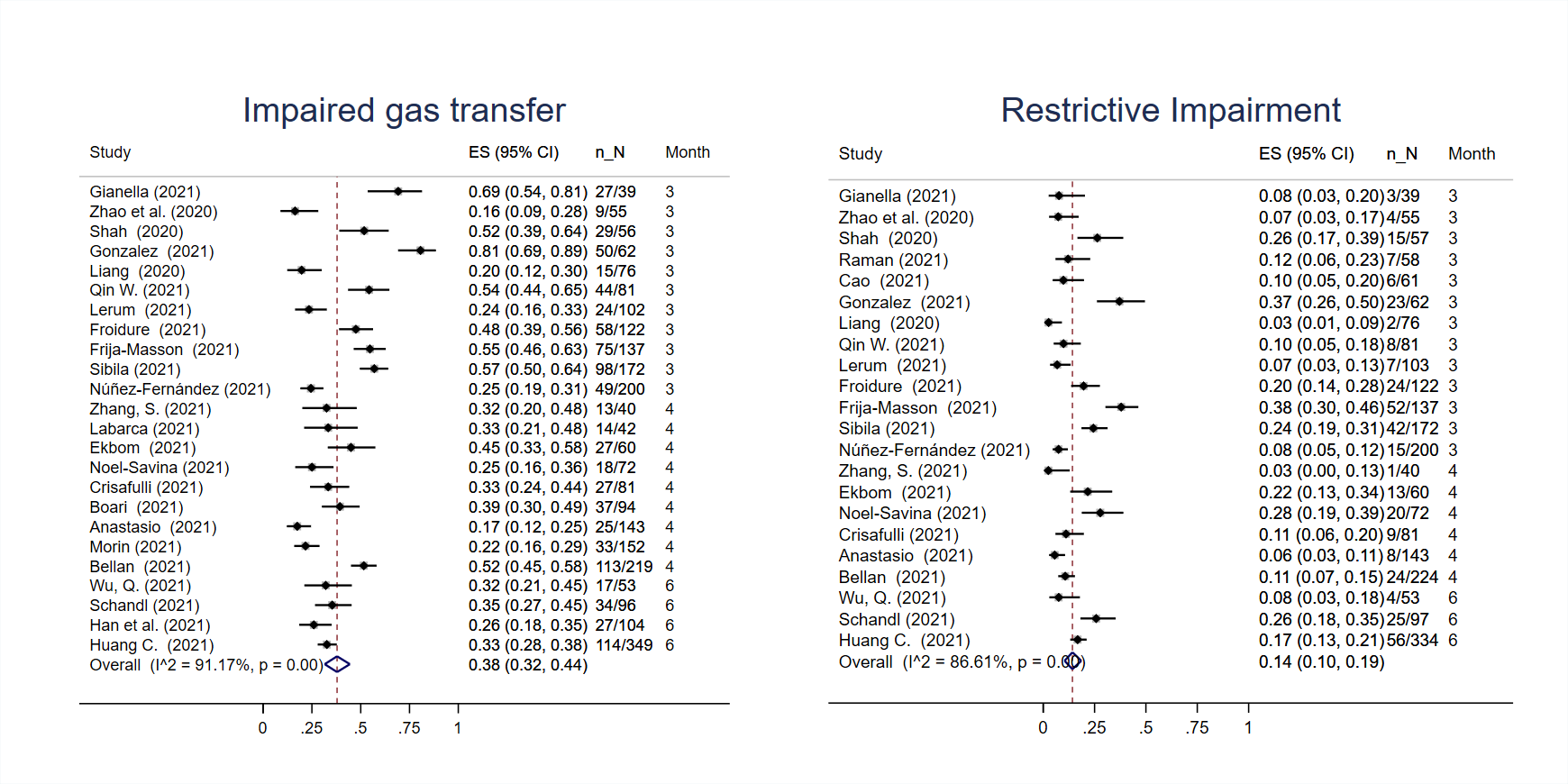

### Supplementary Figure 7. Lung function findings at follow-up in Influenza, SARS-CoV, MERS-CoV studies

Estimates are reported as proportion of number of lung function tests showing the outcome of interest (n) on the total number of exams performed (N) and 95% confidence interval.

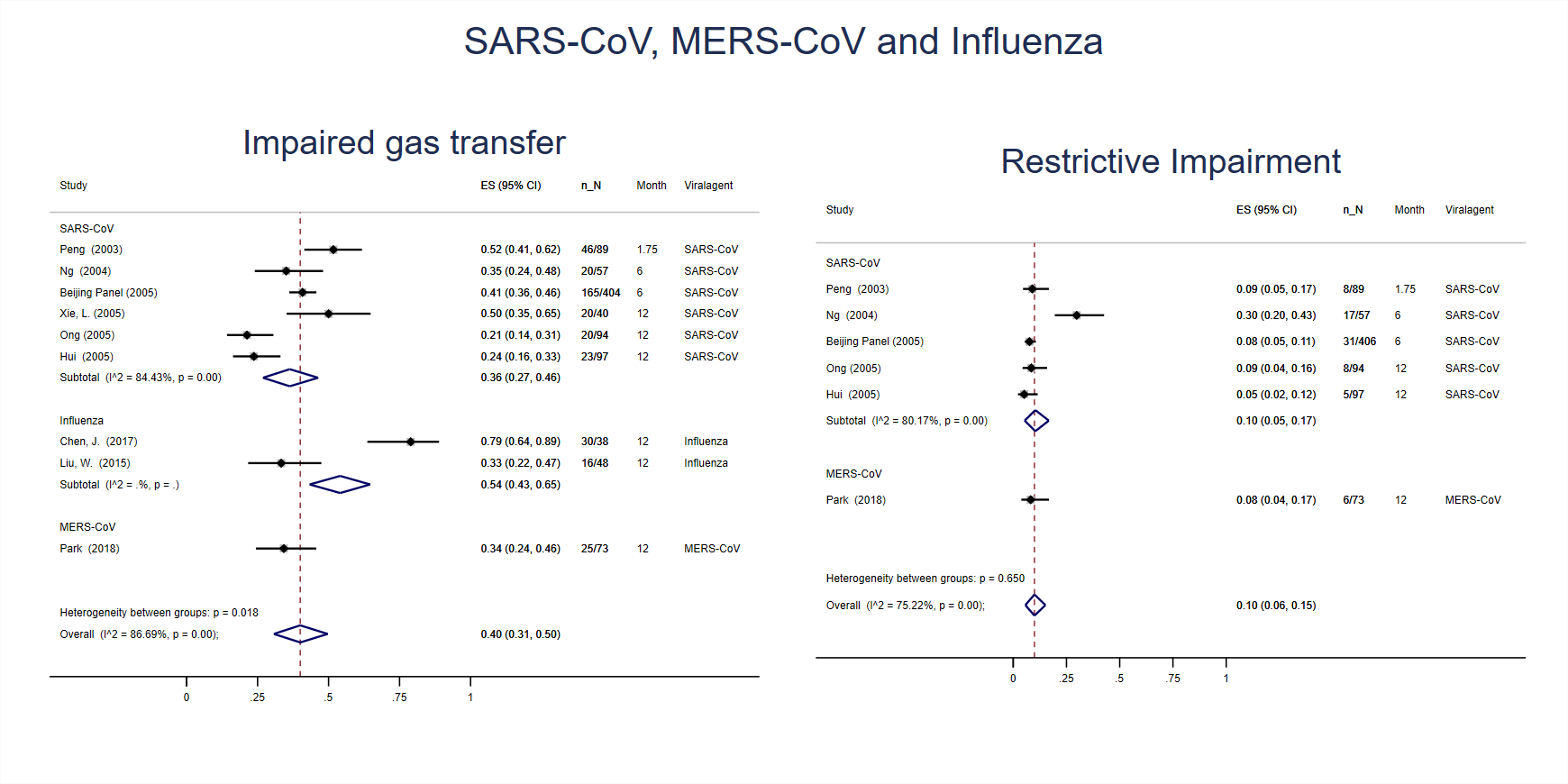

### Supplementary Figure 8. Bubble plots of timing of follow-up in meta regression: radiological sequelae

Timing of follow-up in months. Top panels – inflammatory estimates; bottom panels – fibrotic estimates; left panels – all follow-up, middle panels – sensitivity, right panels – sub analysis.

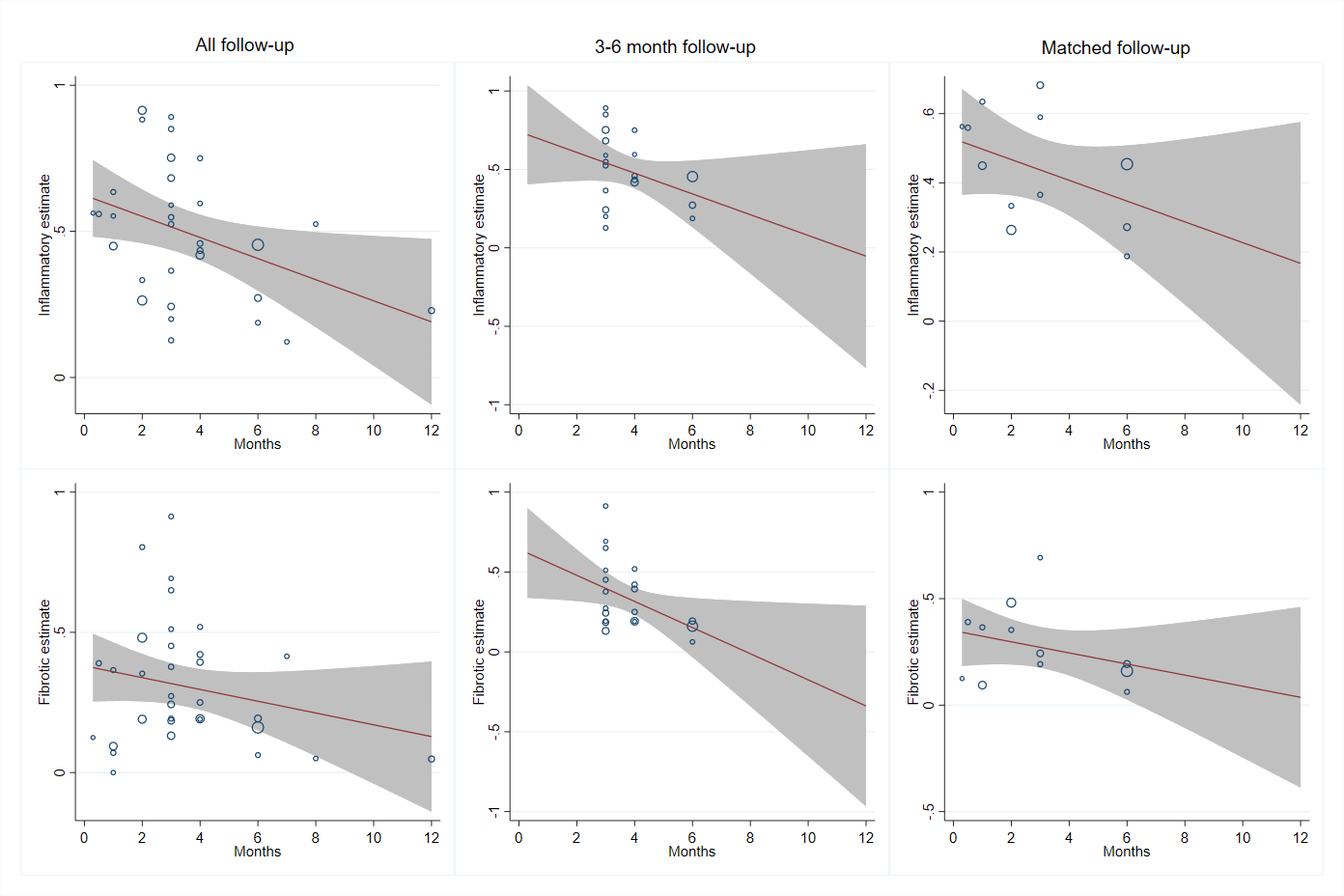

### Supplementary Figure 9. Bubble plots of timing of follow-up in meta regression: physiological sequelae

Timing of follow-up in months. Top panels – impaired gas transfer estimates; bottom panels – restrictive impairment estimates; left panels – all follow-up, right panels – sensitivity.

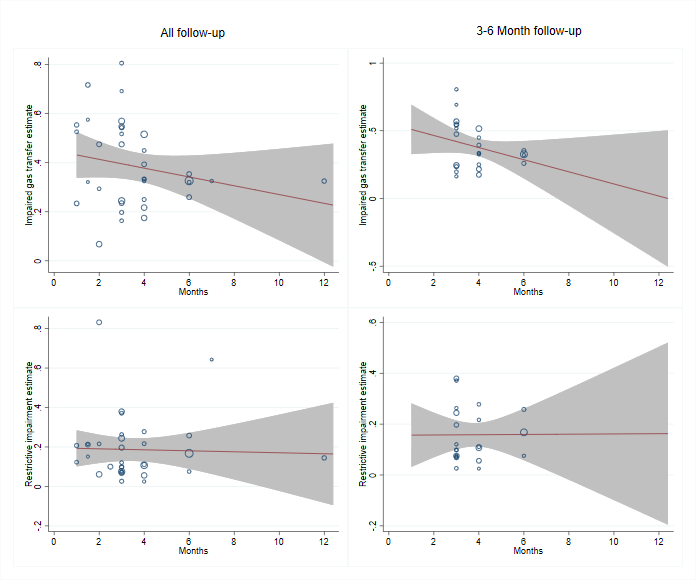

### Supplementary Figure 10. Funnel plots of publication bias: radiological sequelae

Effect size (estimate) plotted with standard error with random effects. A – all follow up, B – sensitivity, C – sub analysis, i – inflammatory, ii – fibrotic.

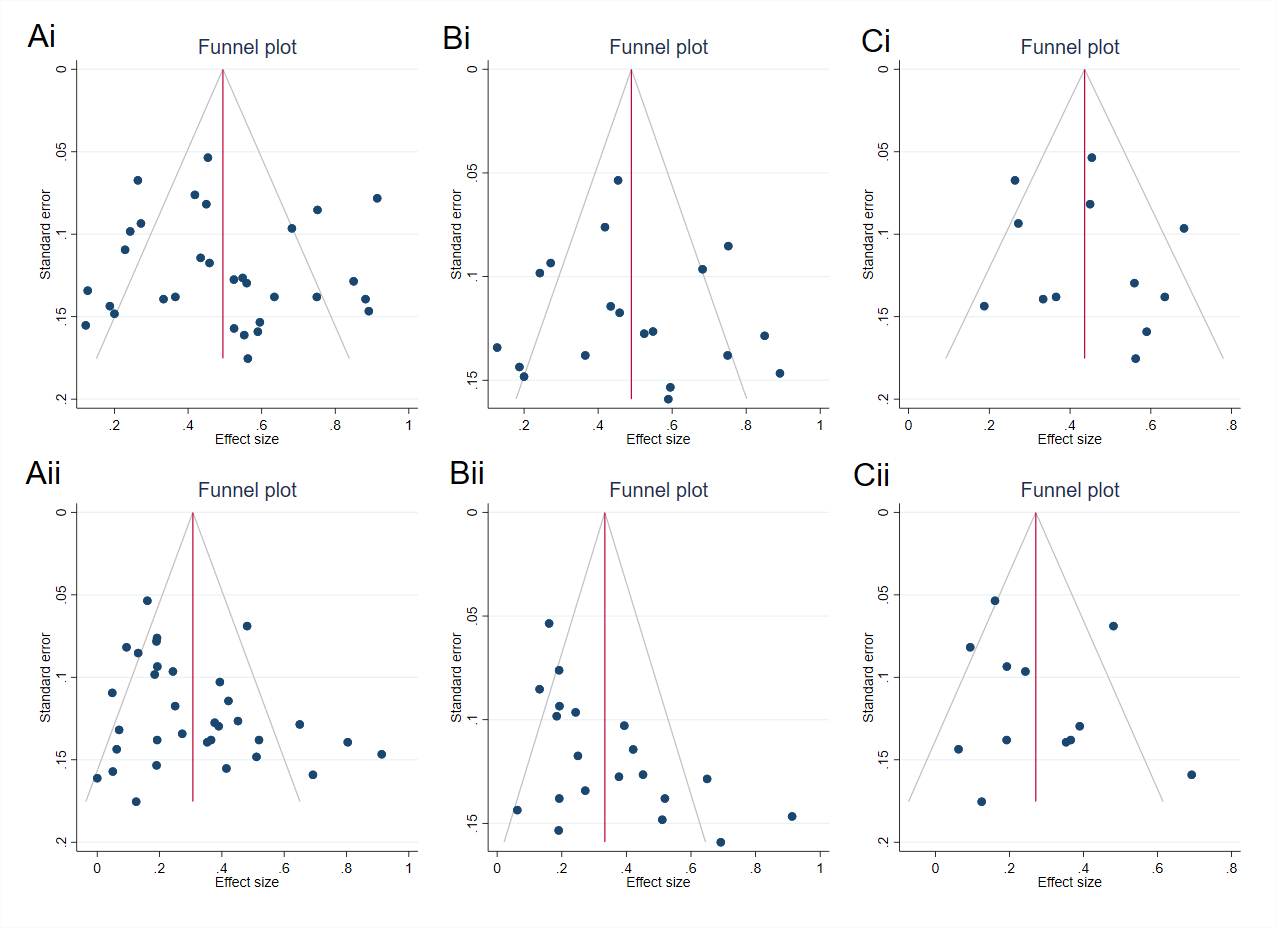

### Supplementary Figure 11. Funnel plots of publication bias: physiological sequelae

Effect size (estimate) plotted with standard error with random effects. A – all follow up, B – sensitivity, i – impaired gas transfer, ii – restrictive impairment.

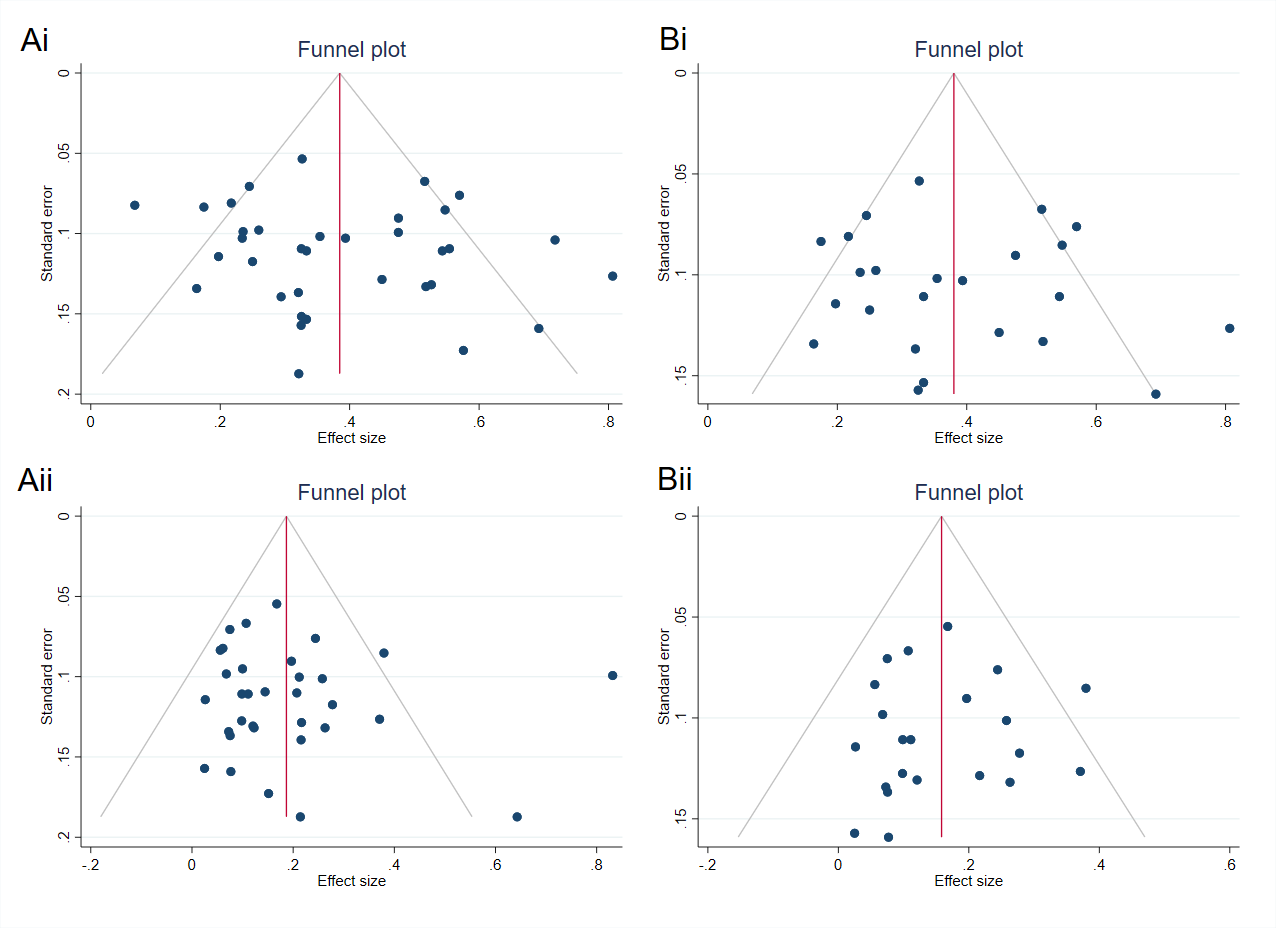

1. 4
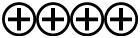
**High** = This research provides a very good indication of the likely effect. The likelihood that the effect will be substantially different** is low.

   3
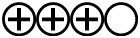
**Moderate** = This research provides a good indication of the likely effect. The likelihood that the effect will be substantially different** is moderate.

   2
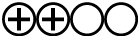
**Low** = This research provides some indication of the likely effect. However, the likelihood that it will be substantially different** is high.

   1
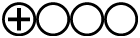
**Very low** = This research does not provide a reliable indication of the likely effect. The likelihood that the effect will be substantially different** is very high.

   ** Substantially different = a large enough difference that it might affect a decision
